# The Effect of the COVID-19 Pandemic on Mental Health in Low and Middle Income Countries

**DOI:** 10.1101/2022.07.29.22278182

**Authors:** Nursena Aksunger, Corey Vernot, Rebecca Littman, Maarten Voors, Niccolo Meriggi, Amanuel Abajobir, Bernd Beber, Katherine Dai, Dennis Egger, Asad Islam, Jocelyn Kelley, Arjun Kharel, Amani Matabaro, Andrés Moya, Pheliciah Mwachofi, Carolyn Nekesa, Eric Ochieng, Tabassum Rahman, Alexandra Scacco, Yvonne van Dalen, Michael Walker, Wendy Janssens, Ahmed Mushfiq Mobarak

## Abstract

We track the effects of the COVID-19 pandemic on mental health in eight Low and Middle Income Countries (LMICs) in Asia, Africa, and South America utilizing repeated surveys of 21,162 individuals. Many respondents were interviewed over multiple rounds pre- and post-pandemic, allowing us to control for time trends and within-year seasonal variation in mental health. We demonstrate how mental health fluctuates with agricultural crop cycles, deteriorating during pre-harvest “lean” periods. Ignoring this seasonal variation leads to unreliable inferences about the effects of the pandemic. Controlling for seasonality, we document a large, significant, negative impact of the pandemic on mental health, especially during the early months of lockdown. In a random effects aggregation across samples, depression symptoms increased by around 0.3 standard deviations in the four months following the onset of the pandemic. The pandemic could leave a lasting legacy of depression. Absent policy interventions, this could have adverse long-term consequences, particularly in settings with limited mental health support services, which is characteristic of many LMICs.

## 1. Introduction

The COVID-19 pandemic and associated containment policies sparked a global economic crisis of unprecedented depth and scale. The adverse effects on living standards were most acutely felt in Low and Middle Income Countries (LMICs) (Egger et al., 2021; Miguel & Mobarak, 2021). A global review primarily focused on high-income country studies reports that the combination of the COVID-19 death toll and the economic downturn increased the prevalence of depression by 27.6% (Santomauro et al., 2021). The virus sparked widespread fear of infection. The stigma of infection generated anxiety, deaths of family members caused anguish, loss of employment and income created economic stress and food insecurity, and mobility restrictions and lockdowns caused separation, loneliness, mental distress, and higher suicidal tendency (Bagcchi, 2020; Brooks et al., 2020; McIntyre & Lee, 2020; Rahman et al., 2021). While the available evidence documenting these effects has focused more on High Income Countries (HICs) (Aknin et al., 2021; Beutel et al., 2021; Daly & Robinson, 2021; Daly et al., 2020; Pierce et al., 2021; Proto & Quintana-Domeque, 2021; Raina et al., 2021; Sun et al., 2021; van der Velden et al., 2020; Varga et al., 2021), these phenomena may be more severe in LMICs. Many LMICs experienced strict containment measures, suffer from poorer access to health care, lower health insurance coverage, shortage of medical supplies (including COVID-19 vaccines), and informal labor markets characterized by a lack of social safety nets and uncertainties created by a lack of relevant information (Vigo et al., 2020).

Due to the limited availability of data, the consequences of the COVID-19 pandemic in LMICs have received less attention in the academic literature and policy dialogues. Limited pre-COVID-19 period data also makes it more challenging to track *changes* in the prevalence of mental health issues in LMICs post-pandemic (Kola et al., 2021; Leucht et al., 2021). During the pandemic, stress may have been more acute in LMICs’ populations due to more profound economic crises that resulted in widespread food insecurity (Egger et al., 2021). An increase in the prevalence of depression could also be more consequential for LMIC societies due to the lack of resources devoted to mental health services and the potential impairment in economic productivity (Herrman et al., 2022). LMICs’ residents account for more than 80% of the years of healthy life lost due to disability (YLD) associated with mental disorders (World Health Organization, 2020a). Still, less than 1.6% of national health budgets are spent on mental health in these countries, with only one psychiatrist serving 200,000+ people on average (United For Global Mental Health, 2020; World Health Organization, 2022).

We make an important discovery in the process of tracking the mental health consequences of the pandemic in LMICs. We observe large within-year fluctuations in mental health indicators driven by agricultural crop cycles, in which mental health deteriorates during pre-harvest “lean” periods and improves after the harvest. Those fluctuations can dwarf pandemic effects and risk conflating seasonality with the pandemic, depending on when COVID-19 had arrived in a country during the local crop cycle. This makes one-time post-COVID-19 outbreak assessments of mental health outcomes more difficult to interpret. A few other studies in agrarian settings have documented links between weather shocks, the time to harvest, and mental health outcomes such as well-being, cognitive function, and fluid intelligence (Chemin et al., 2013; Lichand & Mani, 2019; Mani et al., 2013).

This study assesses changes in depression in LMICs, which is the most prevalent mental disorder among adults (World Health Organization, 2022).In order to avoid misattributing the effect of seasonal fluctuations in mental health to the pandemic, we track populations over multiple rounds, both pre-and post-pandemic, in eight countries from Sub-Saharan Africa (SSA), South Asia, and South America. This allows us to net out season and time trends. In two countries – Nepal and Kenya – we collected high-frequency data on mental health that we can directly match to seasonal poverty and hunger, allowing us to highlight the importance of this adjustment clearly. We find an adverse effect on mental health during the post-COVID-19 period even after netting out the fluctuations due to seasonal effects.

While a few studies have analyzed the mental health impact of COVID-19 using data collected before and after the pandemic in individual middle-income countries (Hamadani et al., 2020; Herrera et al., 2021; Moya et al., 2021), our study is distinctive in that we use several waves of data from large samples, comprising 68,064 interviews of 21,162 unique individuals across eight different low-resource settings. This allows us to adjust for time trends and seasonality explicitly and generate more reliable estimates of the mental health effect of the pandemic. We find that mental health generally worsened in our sample of LMICs, especially early in the pandemic. A random effects aggregation across our samples shows that the index of depression symptoms increased by around 0.3 standard deviations in the four months following the onset of the pandemic.

Our findings corroborate insights from rural Bangladesh and Colombia, which show worsened maternal mental health during the pandemic (Hamadani et al., 2020; Moya et al., 2021). It also supports findings from Bangladesh, which demonstrate that food insecurity increases women’s stress during the pandemic (Rahman et al., 2021), and findings from Chile showing depression and anxiety symptoms have increased among older adults since the onset of COVID-19 (Herrera et al., 2021). Our primary finding is consistent with the results of high-income country studies, including a meta-analysis of sixty-five longitudinal cohort studies that compare mental health before versus during the COVID-19 pandemic (Robinson et al., 2022). It is also in line with findings from a pre-post COVID-19 analysis which used helpline calls in HICs as a proxy for mental well-being (Brülhart et al., 2021). Other individual country studies from Germany (Beutel et al., 2021), United Kingdom (Daly et al., 2020; Pierce et al., 2021; Proto & Quintana-Domeque, 2021), United States (Daly & Robinson, 2021) and Canada (Raina et al., 2021) also demonstrate worsened mental health problems during the COVID-19 pandemic.

Our results stand in contrast to a study conducted in the Netherlands, which found no change in the prevalence of anxiety and depression symptoms (van der Velden et al., 2020), and contradict a recent systematic review of mental health symptoms during the pandemic, which suggests no widespread negative impacts of COVID-19 (Sun et al., 2021). Only 1 out of 36 studies included in the Sun et al. (2021) review were conducted in low-income settings, so our study is notably different from and complementary to theirs. LMIC residents may have experienced the COVID-19 crisis differently than those in wealthier settings. When people are closer to subsistence levels and have more limited access to social safety nets, unemployment benefits, and affordable health care, the pandemic may translate into more adverse consequences for mental health.

### 1.1 Significance of this Research

About 1 billion people currently suffer from mental or neurological disorders worldwide (World Health Organization, 2022). Mental health problems can affect everyone, yet 82% of people living with a mental health disorder reside in LMICs (World Health Organization, 2020a, 2022). Although there are known effective treatments, mental health is under-resourced, with less than US$ 2 annual spending per person globally and US$ 0.25 per person in LMICs (World Health Organization, 2011). More than 75% of people with mental disorders in LMICs never seek treatment from professionals (World Health Organization, 2020b). These statistics show a high disease burden overlaid with low mental health care coverage in LMICs, which may impede escaping from poverty through psychological poverty traps. Several studies point out that while poverty is a risk factor for mental illnesses, these untreated mental health problems, in turn, may deepen poverty through worse cognitive functioning and short-sighted decision-making (Haushofer & Fehr, 2014; Ridley et al., 2020) and may hinder income growth. Therefore the adverse effects of the pandemic may persist well after the COVID-19 situation is controlled, affecting the well-being of future generations, especially in LMICs. Our findings serve as a call to action for policymakers as they develop post-pandemic recovery policies.

## 2. Data and Methods

### 2.1 Sample construction

Our data consist of ten different samples across eight countries - Bangladesh, Colombia, Democratic Republic of the Congo, Kenya, Nepal, Nigeria, Rwanda, and Sierra Leone (denoted BGD, COL, DRC, KEN1, KEN2, KEN3, NPL, NGA, RWA, and SLE in Table A1) - that have a combined population of nearly 525 million. All our samples are from specific sub-populations within each country.

Each sample includes an assessment of depression symptoms collected before and after the onset of the COVID-19 pandemic. For our main analysis, we denote the ‘onset’ as the date when COVID-19 was designated as a pandemic by the World Health Organization (11 March 2020). In supplemental analyses, we vary the date of COVID-19 onset by country and define this as the date of the largest increase in COVID-19 mitigation policy stringency. Information on the exact timing of the first COVID-19 cases, government-imposed social distancing policies, and survey implementation dates for all samples is described in Appendix B in the Supplementary Materials.

Data were collected from the same households in each survey wave. An advantage of this panel structure is that we have pre–COVID-19 baseline data on mental health for all samples. The primary reporting unit for the mental health survey questions is the individual.

### 2.2 Survey methods and timing

In most samples, post–COVID-19 data were collected via telephone interviews to minimize in-person contact and comply with government social distancing guidelines. Interviews were conducted by local enumerators in each country, with procedures to match languages, dialects, and accents between respondents and enumerators. Phone surveys enabled rapid data collection. However, a drawback of the switch to phone-based data collection is that it may lead to selective attrition of low-income families without phones or living in low-connectivity areas. In one sample (KEN3), this was handled by providing a basic feature phone to respondents who did not have one. For the DRC sample, phone surveys were impossible due to the low coverage of mobile phone networks, and data were collected through in-person interviews.

The COVID-19 pandemic and mental health outcomes may show a spurious relationship depending on the calendar time at data collection. In Figure A1, we summarize the pre- and post-COVID-19 survey dates in each sample and relate them to the timing of the lean season and onset of the COVID-19 pandemic. The post–COVID-19 survey wave in seven samples (BGD, KEN1, KEN2, KEN3, NPL, NGA, and RWA) overlaps with the “lean” season when food stocks in the rural population are generally low, and food prices are high. While this decreases households’ capacity to cope with economic shocks, it is also a low agricultural labor demand period in these settings. It may have independent effects on household resilience to unemployment and mental health shocks (Egger et al., 2021). For example, at the outbreak of the pandemic in March 2020, many rural households in Bangladesh, the Democratic Republic of the Congo, and Rwanda were entering the “lean” season. Three samples (COL, KEN3, and RWA) conducted high-frequency surveys spanning a long-enough period to examine the evolution of post-COVID-19 effects over time. In two samples (NPL and KEN2), we can directly compare mental health changes to the typical food security pattern reported in previous years, as described in more detail below.

### 2.3 Construction of main variables

Our primary outcome is an index of symptoms related to depression. Substantively, depression differs from other mental disorders in its ability to affect daily functioning on a very short-term basis, which can translate into a rapid loss of productivity and earnings. Furthermore, analyzing the extent of depression symptoms in LMICs is important because 80% of the years of healthy life lost due to depression disorders occur in these countries (World Health Organization, 2020a). Our samples contained multiple question items related to depression. Table A2 in the Supplementary Materials provides detailed descriptions of how the mental health outcome variables were constructed in each sample. The reference period in all questionnaires was the previous seven days unless otherwise noted in Table A2. We calculated the index in two ways. For the five samples that collected data using a validated mental health screening tool (BGD, COL, NGA, DRC, and KEN3), we followed the prescribed calculation method, i.e., the summed scores of the Center for Epidemiological Studies-Depression (CES-D) scale for BGD and KEN3, the Symptom Checklist-90-R (SCL-90R) in COL, the 15-item Hopkins Symptom Checklist for DRC, and the World Health Organisation-Five Well-Being Index (WHO-5) for NGA, as explained in Table A2. We then reverse-scored and transformed these measures to have a standard deviation equal to 1 across individuals in the first baseline wave of our panel data, with higher scores showing better mental health.

In the remaining samples (KEN1, KEN2, SLE, NPL, RWA), the mental health questions do not fuly come from a standardized tool. In those samples, we used only those common depression-related questions across all surveys. To maximize consistency in reporting results, we constructed indices of the available depression items as follows. First, we standardized the ordinal scales of each depression item to have a mean 0 and a standard deviation 1 in the baseline survey. We then constructed the index as the weighted average of these standardized depression items, using three different weighting methods. Our preferred measure uses validated items where available and weights all items equally in samples without validated measures. The second measure uses the first factor loadings from a factor analysis as weights. This measure places more weight on strongly correlated items with the goal of identifying the single most important latent variable. The third measure is an inverse-covariance-weighted index calculated as in Anderson (2008). This measure places less weight on strongly correlated items to capture variation in many potential latent variables. We report the results for the validated measures and unweighted indices when discussing the individual samples and the aggregated results. The results for the other weighting methods are available in Table A4 and Table A3 in the Supplementary Materials. Our choice of mental health measure does not significantly change the results.

In the Figure A2, we also plot levels of the depression index over time and include the “COVID-19 case counts” and a measure of the “COVID-19 stringency index” for reference. Data are retrieved from World Health Organization and ourworldindata.org data sources, respectively. The COVID-19 stringency index is a composite metric based on nine policy indicators, such as school closures, workplace closures, and travel bans, re-scaled to range 0 and 100 (100 being the most stringent). We use the stringency index at the national level for all samples.

For two samples where we conduct a more intensive examination of seasonality, we create a food insecurity index by summing standardized items related to whether one or more household members skipped meals or reduced portion size or diet quality in the past seven days (KEN2) or fourteen days (NPL).

### 2.4 Empirical methodology

A straightforward and the most commonly used strategy to investigate the mental health impacts of COVID-19 would be to compare average mental health outcomes for the same individuals at pre- and post-pandemic periods. An implicit assumption in such comparisons is that average mental health levels are not changing over time in the absence of the pandemic. However, there is evidence that mental health indicators fluctuate with agricultural seasons and other events affecting the broader economy (Chemin et al., 2013; Christian et al., 2019; Haushofer & Fehr, 2014). In our own data, we see fluctuating pre-COVID-19 trends in mental health in five of the six samples for which we have multiple pre-COVID-19 survey waves. To assess changes due to the COVID-19 pandemic, we therefore prefer strategies that adjust for non-COVID-19 related trends in mental health, as much as each dataset will allow. Some, but not all, of our samples have data that allow us to adjust for these non-COVID-19 trends directly. Wherever possible, we make these adjustments. In samples where we do not have the data necessary to make adjustments directly, we estimate simpler models with the best possible indirect controls for trends in mental health status unrelated to the pandemic. These strategies are described in detail below. This approach implies that our analysis will combine different estimation strategies for different samples depending on the data structure in each sample.

An alternative would be to ignore non-COVID-19 trends altogether and estimate the simpler model without adjustments in all samples. While this alternative would have the benefits of simplicity and consistency, we prefer estimates that use what we believe is the best estimation strategy in each sample, given the scope of the data we have. Nevertheless, we also report results in Figure A3 where we estimate the same model in every sample without these adjustments. This exercise produces qualitatively similar insights in the aggregate, but the coefficients are less precisely estimated.

Although we use different preferred models in different samples, we still estimate a single average effect of the pandemic by combining our estimates from different samples as one would in a meta-analysis. We do this using a random effects model, the standard tool for meta-analysis, where we allow the true effect to vary across different studies.

#### Estimation Strategies Used

We control for both month and year trends in our estimation in the most extended samples with multiple survey waves pre-COVID-19 spanning a full year or more. Specifically, we estimate:

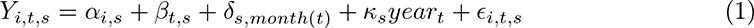

where *Y_i,t,s_* is the outcome depression of individual *i* at time *t* in sample *s*, and *α_i,s_* captures individual fixed effects. *t* = 0 during the pre-COVID period and takes on integer values for discrete time-periods in the post-COVID period. The entire pre-COVID-19 period is our omitted category in all specifications (*β*_0*,s*_ = 0) implying that *β_t,s_* is the difference between mental health during the pre-COVID period and period *t* in sample *s*.

Depending on the granularity of *t*, we can have multiple post-COVID-19 time periods, allowing for different impacts on mental health earlier vs. later in the pandemic. When we aggregate results across samples, we divide the post-COVID-19 period into two time periods: 0-4 months post-COVID-19 and more than 4 months post-COVID-19. We choose this relatively small number of post-COVID-19 time periods to allow us to aggregate across many samples with different timings of surveys, while still allowing for some dynamics in the effects on mental health throughout the pandemic. When estimating models for individual samples, we use more granular time periods post-COVID-19, defining time periods of 0-2, 2-4, 4-6, 6-9, 9-12, and 12-15 months post-COVID-19.

In our samples with multiple years of pre-COVID-19 data (RWA, COL, KEN1), we allow for both seasonal trends and year-on-year trends in mental health in our model as in Equation 1. Here, *δ_s,month_*_(*t*)_ is a month-of-year fixed-effect and *κ_s_* is the year-on-year increase in mental health prior to the pandemic.

In our samples with multiple waves of pre-COVID-19 data spanning less than a full calendar year, but with supplemental data on typical seasonal food security (KEN2 and NPL), we estimate Equation 2, in which we control for the average level of our food insecurity index in respective months pre-COVID-19, designated as *x_t,s_*, as follows:

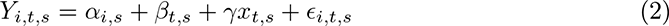

where *β_t,s_* is zero during all pre-COVID-19 time periods, as in Equation 1.

In the KEN2 sample, pre-COVID-19 mental health data were collected three years prior to the pandemic. This large gap in time between the pre- and post-COVID-19 data collection could exacerbate bias from secular trends in mental health over time; if mental health is changing substantially from year to year, the more time in between our pre-COVID-19 data and the pandemic, the worse of a counterfactual that pre-data will be. However, the KEN2 sample collected data on food security over full calendar years that spanned both 2014-2015 and 2016-2017. This allows us to estimate and incorporate any secular trends in food security into our control variables. This is reflected in Equation 3:

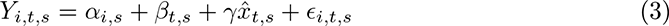

where *x̂_t,s_* is estimated from the 2014-15 and 2016-17 data on food security with both month-of-year and linear year-on-year time trends, as in Equation 4:

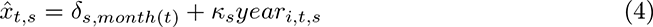

Here, *year_i,t,s_* is a continuous variable indicating the date the survey was conducted and transformed into yearly units for interpretation. During the pre-COVID-19 period, the value is set to the exact date for each survey, so that we can use this variation to estimate *κ_s_*. During the post-COVID-19 period, the value of *year_i,t,s_* is set to the average survey date of all respondents in discrete time period *t* in sample *s*. This is done so that *year_i,t,s_* will be intentionally colinear with our indicators for discrete time periods during the post-COVID-19 period, so only variation from the pre-COVID-19 period will be used to estimate *κ_s_*.

For one sample (KEN3), we have multiple waves of pre-COVID-19 data spanning less than a full year and we do not have supplemental data on typical seasonal food security. We choose to control for linear time trends in this sample. In the remaining samples with only one wave of pre-COVID-19 data (BGD, DRC, NGA, SLE), we regress mental health on individual fixed-effects and the pre-post-COVID-19 time period categories only.

To estimate the average effects of the pandemic on mental health across our samples, we aggregate the estimates across countries using a random effects model and estimate the inter-sample variance in effects *τ* ^2^ using restricted maximum likelihood (following recent simulation studies by Langan et al. (2019), Petropoulou and Mavridis (2017), and Viechtbauer (2005)).

For figures showing trends in mental health for individual samples, we break post-COVID-19 survey waves into 1-month intervals^1^ and plot the average survey date during the time period on the x-axis.

## 3. Results

### 3.1 Seasonal Trends in Mental Health

We start with a deeper investigation of seasonal trends in depression. Figure 1 zooms in two of our samples (NPL and KEN2), for which we observe mental health across multiple years. We have less than a full year of pre-COVID-19 data in these samples, so we cannot adjust for independent year-on-year improvements in mental health. However, we do have detailed pre-COVID-19 data on typical trends in seasonal food insecurity in these samples, which we can use to adjust for recurring agricultural seasons.

**Figure 1:**
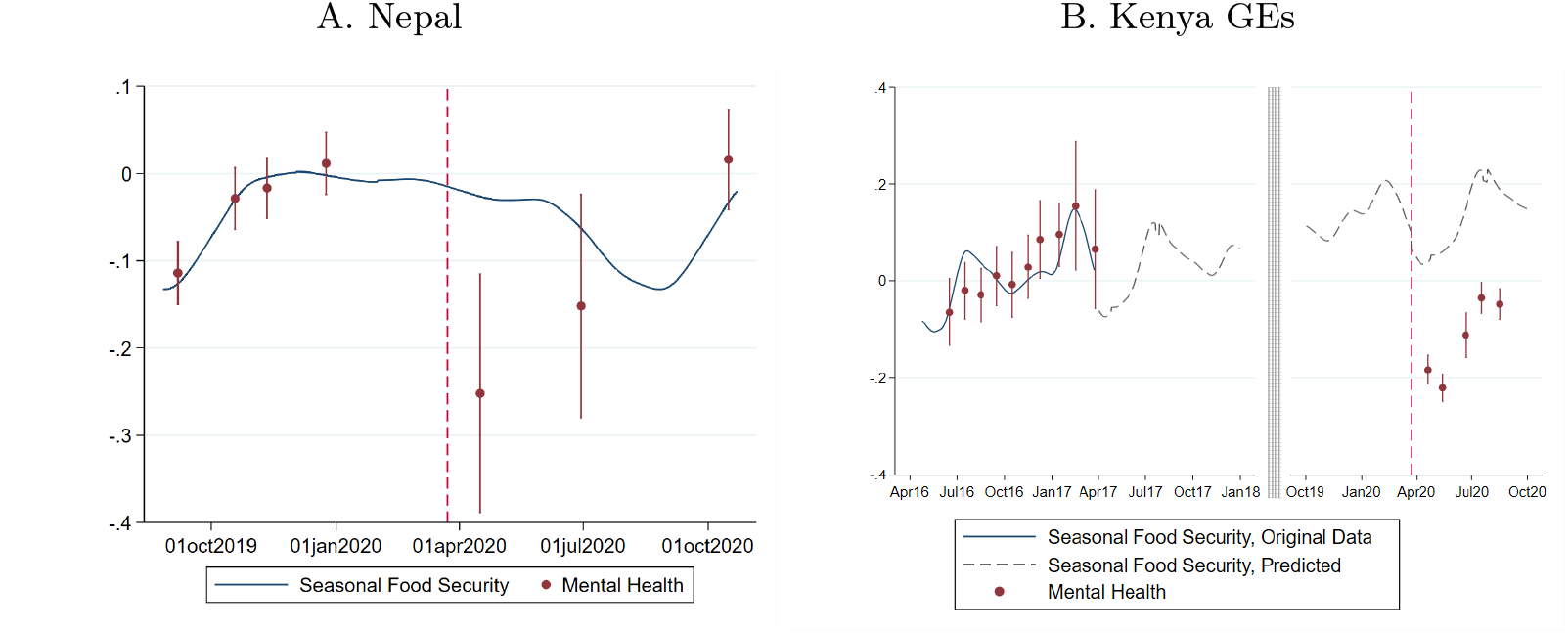
Seasonal Food Security and Pre-Trends. Note: Figure shows evolution of seasonal food security (indicated by continuous blue line) and in mental health (indicated by red points and error bars) over time before and after the onset of the pandemic (indicated by the red vertical dashed line). The line for food security is the predicted values from a lowess model using data from an index of food security items. Panel A shows data from our NPL sample. Panel B shows data from our KEN2 sample. In panel B, the solid blue line shows the 2016-17 period during which we have both mental health data and food security data that we used to estimate the lowess. The dashed line is our extrapolated prediction based on this data. Not pictured are the 2014-15 period during which we have food security data but no mental health data. The red dots and vertical lines are predicted values and associated 95% confidence intervals for our positively coded depression index. The predicted values are based on discrete 1-month intervals using coefficients from Equation 1. The x-axis value for each dot and confidence interval is the average date of surveys conducted during that time interval.

In both samples, we see clear evidence of a positive trend in mental health status prior to the pandemic, closely following the fluctuations in food security (comparing the blue line and the red dots in Figure 1). We also see that mental health in April 2020 is well below the pre-COVID-19 average and improves substantially from April to October 2020. However, it is difficult to interpret these post-COVID-19 fluctuations without information on seasonal trends. Does the pandemic lead to short-term negative mental health impacts, which gradually reduce over time? Or are these post-COVID-19 improvements in mental health due to normal seasonal dynamics?

By adding data on typical seasonal trends in food security, we can paint a clearer picture of the mental health consequences of the COVID-19 pandemic. In NPL, the “lean” season occurs roughly from July to September, and we see little difference between food security in April and October of a typical year. This suggests that the pandemic had an immediate negative effect on mental health in April, which faded over time. In contrast, in KEN2, food security rises substantially in a typical year from April to October. Moreover, the 2014-15 and 2016-17 data on food security show an independent average increase of 0.06 SD per year in the food security index. We therefore incorporate this year-on-year trend into our measure of seasonal food security. The dotted blue line in panel B of Figure 2 shows the extrapolated average of seasonal food security in 2014-15 and 2016-17 in the sample, plus a linear time trend of 0.06 SD per year. Clearly, we cannot attribute all of the improved mental health from April 2020 to October 2020 to fading effects of the pandemic; seasonal effects contribute to the improvement as well.

**Figure 2:**
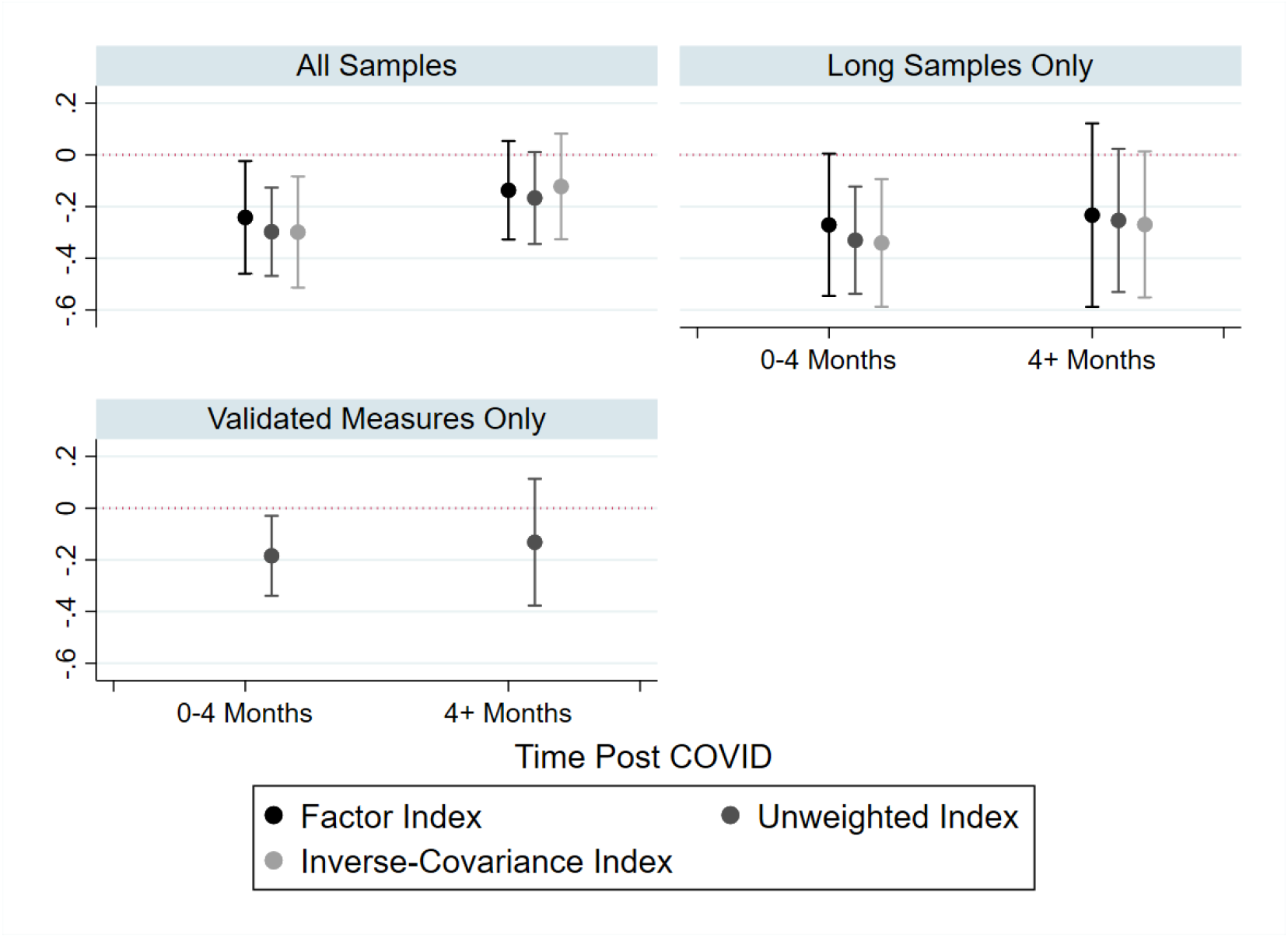
Random Effects Aggregates Different Estimation Strategies. Notes. The figure plots the average change in mental health pre-post COVID across samples. The Y-axis is the change in mental health relative to pre-COVID. The X-axis shows the time after the onset of COVID-19 discretized into two groups: 0-4 months, and more than 4 months. Point estimates are represented as dots with 95% confidence intervals as whiskers. The color of the points indicates which method of weighting our indices was used: the factor loadings (black, left), the unweighted index (dark gray, middle) or the inverse-covariance weights (light gray, right). The three panels use three different sets of samples in the estimates. The first panel includes all samples. The second panel (top right) uses only samples with over a year of pre-COVID-19 mental health data (RWA, COL, KEN1) or supplemental data on typical seasonality in welfare (KEN2, NPL). The third panel (bottom left) uses only samples with validated depression measures (BGD, COL, NGA, DRC, KEN2). The average effect sizes and confidence intervals are estimated from a random-effects model using restricted maximum likelihood to estimate the variance in true effects across samples. The estimates range from a min of −0.34 SD (the inverse covariance weighted index, 0-4 months post-COVID-19, using samples with long panels or seasonality data) to a max of −0.08 SD (inverse covariance weighted index, 4+ months post-COVID, samples with validated measures only).

### 3.2 Mental Health Effects of COVID-19

The aggregate (cross-country) estimates of the impact of the COVID-19 pandemic on mental health are shown in Figure 2. The two aggregate post-COVID-19 periods (0-4 months post-COVID-19 and more than 4 months post-COVID-19) are shown on the x-axis of each panel. The three panels of Figure 2 vary in the set of samples that are included - the top-left panel include all samples, the top-right panel includes only samples with either multi-year pre-COVID-19 waves or seasonal food security data, and the bottom-left panel includes only samples for which depression was measured with a complete validated mental health screening tool. In the top panels, we show results for three methods of constructing the depression index for those samples without a full, validated scale: a factor analysis which places more weight on positively correlated mental health questions; an unweighted average of mental health items (our preferred method); and the inverse-covariance weighted index of Anderson (2008).

Our short-run aggregates (0-4 months post-COVID) make use of the six samples where we have survey waves within four months of the pandemic: COL, KEN1, KEN2, KEN3, NPL, and RWA, further adjusted to the inclusion criteria per panel in Figure 2.^2^ The longer-run aggregates make use of nine samples with survey waves 4 months or more after the onset of the pandemic: BGD, COL, DRC, KEN1, KEN2, NGA, NPL, SLE, and RWA.^3^ The longer-run aggregates weight each sample more equally than the short-run aggregates because the heterogeneity in effects across samples is larger 4+ months after the onset of the pandemic.

Overall, Figure 2 shows high consistency in our aggregate results across inclusion criteria in the three panels and across methods of constructing the depression index. In the short run, between 0 and 4 months after the onset of the pandemic, we see a decline in depression measures of between 0.27 and 0.34 standard deviations. Over the longer run, 4 months or more post-COVID-19, evidence suggests mental health status rebound somewhat. However, the improvement is small and the aggregate estimates remain negative, albeit not significantly different from zero. In our five samples with long pre-panels or seasonality data where we have both short-term and longer-term estimates (COL, KEN1, RWA, KEN2, NPL), mental health improves on average by 0.08 SD (95% confidence interval (CI) [-0.07, 0.22]) from 0-4 months to 4+ months.

Table 1 shows the full results for each of the ten samples. Our dependent variable is the un-weighted depression index (see the results for the factor analysis as well as inverse-covariance weighted index in Table A3 and Table A4. Results are qualitatively similar.). The table shows the samples where we adjust for seasonal effects and time trends in the first five columns. Specifically, columns 1-3 are based on the full specification in Equation 1, and columns 4 and 5 are based on the models that control for seasonal food security as in Equation 2 and Equation 3-Equation 4, respectively. The remaining five samples with less than one year of pre-COVID data and no supplemental data on food security trends are in columns 6-10.

**Table 1:**
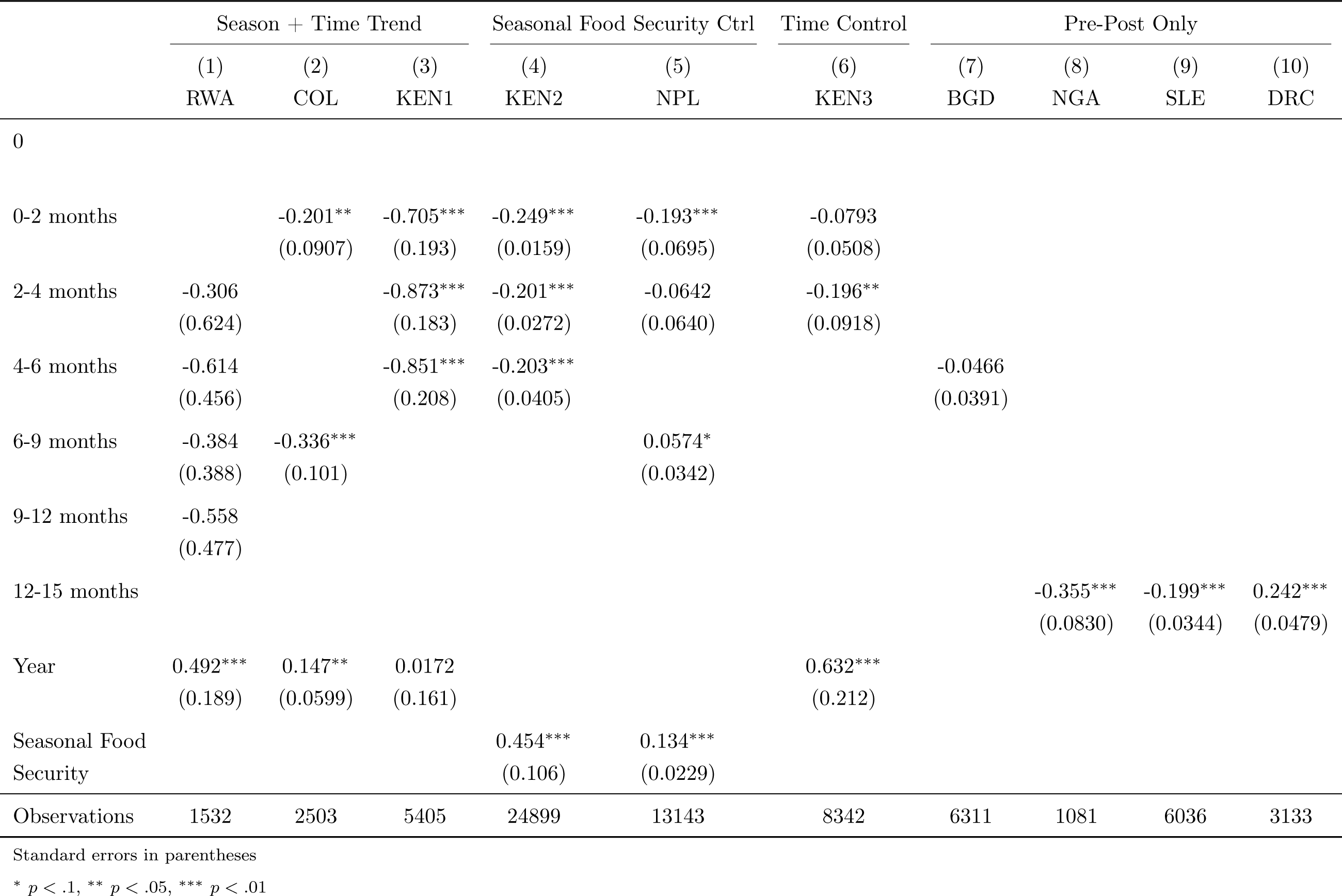
Best Estimates from Each Sample.

The results are consistent with Figure 2 for most samples. COL, KEN1, KEN2, KEN3, NGA and SLE all show negative and significant coefficients throughout the survey waves implying a long-lasting impact of COVID-19 on mental health. For RWA, the coefficients are negative and about −0.3 SD but not significant, largely due to increased noise in the estimation. NPL and DRC show a reverse in sign over the longer run. For NPL, this likely reflects that mental health status returns to a pre-COVID-19 mean as suggested in Fig ure 1A, where we can’t separate the observed mental health levels from the seasonal trend. For DRC, similar explanations hold. The interpretation is more speculative, as we lack data on seasonal variation in mental health.

### 3.3 Alternative Specification: Regression Discontinuity

Our specifications in columns 1-3 and 6 of Table 1 relying on the assumption that pre-pandemic trends continue linearly in the post-pandemic period. However, this assumption may be too strong, especially when we use a relatively short period of pre-COVID-19 data to estimate counterfactual mental health status long into the pandemic (as is the case for the KEN2 sample). Instead, a weaker assumption is that independent time trends in mental health outcomes are locally linear around the onset of the pandemic. With this assumption, we can estimate the very short-run impact of the pandemic by comparing mental health outcomes just before and just after the onset of the pandemic. This approach of comparing outcomes before and after a specific threshold is known as regression discontinuity (RD) design. For a survey of regression discontinuity applications in economics, see Lee and Lemieux (2010). We estimate the RD specification around the initial onset of COVID-19 in the five samples where we have multiple pre- and post-COVID-19 survey waves within 3 months of the start of the pandemic.

The RD estimates presented in Table 2 for the NPL, COL and KEN1 samples are similar to our estimates of the impact 0-2 months post-pandemic in columns 2, 3 and 5 of Table 1. The RD estimate for KEN3 is effectively zero and insignificant. This is roughly in line with the relatively small coefficient in row 1, column (6) of Table 1. In contrast, the RD estimate for RWA is positive and significantly different from our estimate in Table 1. One reason for this difference is that our estimates in Table 1 include month-of-year fixed effects, effectively comparing 2020 data to the same months in 2019. The months in which survey waves occur post-COVID-19 in RWA are relatively high mental-health months in terms of seasonality as deduced from pre-COVID-19 fluctuations. Another reason for the difference is that Table 1 assumes pre-COVID-19 trends continue post-COVID-19 while the RD specification estimates separate slopes pre- and post-COVID-19. This is easiest to visualize in the figure in column (6) of Table 2. Overall, the magnitude of the aggregate for these five countries decreases by around a third and is no longer statistically significant (p = 0.18)

**Table 2:**
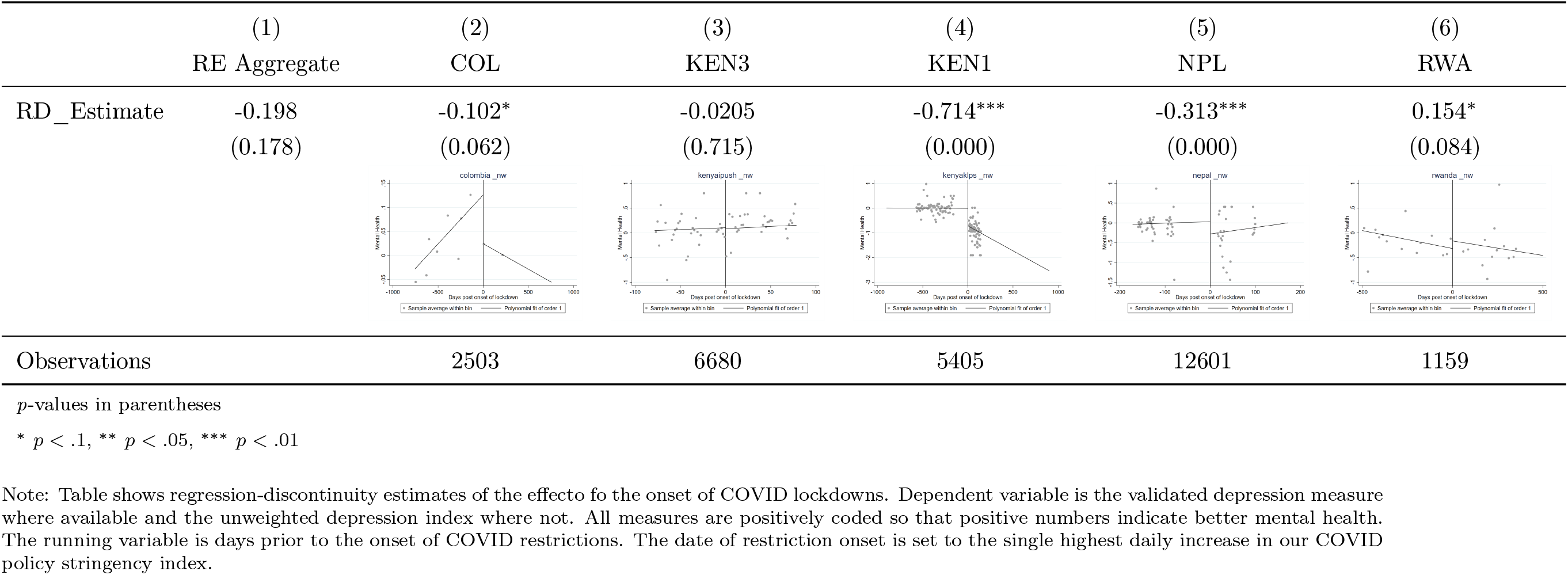
RD Estimates from Samples With Surveys Near Pandemic Onset.

|  | (1)<br>RE Aggregate | (2)<br>COL | (3)<br>KEN3 | (4)<br>KEN1 | (5)<br>NPL | (6)<br>RWA |
| --- | --- | --- | --- | --- | --- | --- |
| RD_Estimate | -0.198<br>(0.178) | -0.102*<br>(0.062) | -0.0205<br>(0.715) | -0.714***<br>(0.000) | -0.313***<br>(0.000) | 0.154*<br>(0.084) |
| Observations |  | 2503 | 6680 | 5405 | 12601 | 1159 |

*p*-values in parentheses
\* $p < .1$ , \*\* $p < .05$ , \*\*\* $p < .01$
Note: Table shows regression-discontinuity estimates of the effect to the onset of COVID lockdowns. Dependent variable is the validated depression measure where available and the unweighted depression index where not. All measures are positively coded so that positive numbers indicate better mental health. The running variable is days prior to the onset of COVID restrictions. The date of restriction onset is set to the single highest daily increase in our COVID policy stringency index.

## 4. Conclusions and Implications for Policy

We interviewed more than 20,000 individuals from eight LMICs in Asia, Sub-Saharan Africa and South America both before and after the onset of the COVID-19 pandemic. We carefully estimate the effect of the pandemic on mental health using multiple estimation strategies that account for plausible threats to inference. Taken together, this analysis paints a clear, consistent, and robust picture: the pandemic produced a substantively and statistically significant increase in depression in LMICs, even after netting out seasonal fluctuations and time trends.

Existing multi-country studies of the mental health consequences of the pandemic largely focus on high-income country samples, and our study, therefore, makes an important contribution to this literature. Populations in LMICs have been at a higher risk for mental health problems resulting from the COVID-19 pandemic, both because the pandemic led to considerably more severe economic crises in these settings (Miguel & Mobarak, 2021), and because mental health support services are sparse. A random-effects aggregation across samples shows that depression symptoms increased by around 0.3 standard deviations in the four months following the onset of the pandemic. Over the longer run, we see some signs of recovery, but in our long panels, where we control for seasonal effects, we do not see a clear sign of mental health fully bouncing back to pre-pandemic levels.

Our LMIC-focused investigation also produces an empirical insight that has important methodological implications for the work of other researchers and practitioners interested in tracking mental health in low-income settings. In agrarian societies, mental health fluctuates within the year, in line with periods of scarcity versus plenty, before versus after crop harvests. Ignoring seasonal fluctuations can lead to spurious associations between other events (like a pandemic) and mental health, and bias estimates. Addressing the effects of seasonality properly requires multiple rounds of high-frequency data collected from a stable sample of individuals.

Mental health disorders have long been recognized as one of the greatest health burdens globally, with severe repercussions for social welfare (Arango et al., 2018; Wykes et al., 2015). Poorer people often bear the greater burden of mental disorders, both because they are at higher risk due to constant exposure to stressful events, and more limited access to treatment. COVID-19 has exacerbated the existing mental health crisis and exposed the weaknesses of existing health systems (Chang et al., 2021; Choi et al., 2020). Learning from this crisis, it is necessary to rethink how to best address mental health issues in public health strategies. Our results suggest that this has become even more important now, as the pandemic may leave a lasting legacy of depression that affects long-run health and productivity even after the virus subsides. LMIC governments must invest in mental healthcare now.

Global resources allocated to mental health are currently limited. This situation is aggravated in LMICs, where the mental health workforce and essential psychotropics are scarcer (World Health Organization, 2011) and where discussions of mental health problems are met with the social stigma (World Health Organization, 2022). Innovative strategies are required to address this crisis in the absence of a large number of trained mental health care providers. Recent evidence shows that psychotherapy delivered by non-specialists is a cost-effective treatment for depression even after 5 years (Bhat et al., 2022; Patel, 2022). Considering the enormous returns from engaging non-specialists in this sector, policymakers could prioritize task-sharing in mental health care. Frugal solutions like the training of paraprofessionals, lay health workers, and community-based care providers would be a great source of support in increasing the accessibility and affordability of mental health services and such supports can be included in universal healthcare coverage (UHC) (Patel, 2022).

## Data Availability

The data underlying the results presented in the study are available upon request for researchers who meet the criteria for access to confidential data.

## List of Abbreviations

BGD: Bangladesh
CES-D: The Center for Epidemiological Studies-Depression
CI: Confidence interval
COL: Colombia
DRC: Democratic Republic of the Congo
HICs: High Income Countries
KEN: Kenya
LMICs: Low and Middle Income Countries
NPL: Nepal
NGA: Nigeria
YLD: Years of healthy life lost due to disability
RWA: Rwanda
SCL-90R: The Symptom Checklist-90-R
SLE: Sierra Leone
SSA: Sub-Saharan Africa
UHC: Universal care coverage
WHO-5: The World Health Organisation-Five Well-Being Index

## Acknowledgements

We are indebted to study participants for generously giving their time. We are grateful to the staff of Yale Research Initiative on Innovation and Scale (Y-RISE); Centre for the Study of Labour and Mobility in Nepal (CESLAM); The Amsterdam Institute for Global Health and Development (AIGHD), Richard de Groot, Menno Pradhan; Africa Population and Health Research Center (APHRC), Estelle Sidze, Caroline Wainaina; 100WEEKS Director Jeroen de Lange and Rwanda Country Director Gervais Nkurunziza; Semillas de Apego team: Arturo Harker, Blasina Niño, Vilma Reyes, Alicia Lieberman, María José Torres and Juliana Sánchez; IPA Nigeria’s Country Director, Emeka Eluemunor and IPA’s Regional Director for West Africa, Claudia Casarotto; the Sierra Leone Ministry of Energy and Madison Levine, Joseph Levine, Vasudha Ramakrishna; Sarah Khan, Morgan Holmes, the late Jean Paul Zibika, Amani Matabaro Tom and the team at Forcier Consulting, Search for Common Ground, the IMA World Health and funded by the USAID; Innovations for Poverty Action Kenya, Eric Ochieng, Ronald Malaki, Michelle Layvant, Somara Sobharwal, Pooja Suri and Eve Zhang; Vyxer Remit Kenya, Carol Nekesa, Andrew Wabwire, Layna Lowe, Anya Marchenko, Gwyneth Miner, Carlos Paramo, Tilman Graff and Magdalena Larreboure; BRAC Institute of Governance and Development, BRAC Institute of Educational Development, Monash University Australia.

Data collection and research time in this study were supported by grants from Saving Brains–Grand Challenges Canada (grant number SB-POC-1809-19901), Fundación Éxito, Fundación FEMSA, United Way Colombia, and Universidad de los Andes; IPA Peace and Recovery Full Study grant and implemented by IPA Nigeria; the Applied Research Programme on Energy for Economic Growth (EEG) led by Oxford Policy Management (funded by the UK Government through UK Aid), the Dutch Research Council, and the United Nations Office for Project Services (UNOPS); IPA’s Peace and Recovery Initiative, IZA’s G2LMLIC, Berkeley Population Center, NIH and GiveWell; USAID via NORC at the University of Chicago; BRAC and LEGO Foundation (for the HPL program in Bangladesh); International Growth Centre, PEDL, NSF and Weiss Family Fund; the Ministry of National Education of Turkey; Sint Antonius Stichting; and the Joep Lange Institute.

The studies BGD, COL, DRC, KEN1, KEN2, KEN3, NPL, NGA, RWA, and SLE received Institutional Review Board (IRB) approval from BRAC University (ref no. 2019-028-ER); the Ethics Committee of the Universidad de los Andes, Colombia (protocol 786, 2017); the NORC Institutional Review Board (IRB00000967); UC Berkeley, Maseno University; AMREF Ethics and Scientific Review Committee Kenya (P679/2019); Yale University (IRB Protocol 2000025621); IPA (IRB Protocol 15057); Vrije University Amsterdam (20210413.1); and Sierra Leone Ethics and Scientific Review Committee (SLERC 2904202) and Wageningen University (24062020), respectively as denoted in Appendix C.

## Authorship

N.A., C.V., R.L., M.V. and N.M. are co-first authors. W.J. and A.M.M. are co-last authors. A.M.M. is the corresponding author. W.J. and A.M.M. conceived the study, acquired the datasets and provided overall guidance. N.A. led the literature review. N.M., A.A., A.K., A.Moya, A.Matabora, P.M., C.N., M.V. E.O. and Y.v.D. oversaw data collection as part of other research efforts. N.A., C.V., and A.M.M. coordinated the project across study samples. The following verified the underlying data for individual study samples: A.I. and T.R. (Bangladesh), A.Moya (Colombia), M.V., J.K. and A. Matabaro (Democratic Republic of the Congo), E.O. and M.W. (Kenya sample 1), D.E., C.N. and M.W. (Kenya sample 2), N.A., A.A. and W.J. (Kenya sample 3), C.V., A.M.M. and A.K. (Nepal), B.B., P.M. and A.S. (Nigeria), Y.v.D. and W.J. (Rwanda) and N.M., M.V. and A.M.M. (Sierra Leone). N.A. and C.V. collated and processed all datasets used for the analysis and provided the overall replication/reproducibility of results and other research outputs. K.D. processed the additional external data required for the analysis. N.A., C.V., R.L., N.M., W.J. M.V and A.M.M. carried out data interpretation and verified final datasets and analysis. C.V. performed the data analysis. N.A. and C.V. produced output figures with input from M.V., N.M., W.J. and A.M.M. N.A., C.V., M.V., N.M., W.J. and A.M.M. wrote the first draft of the manuscript. All authors read and approved the final version of the manuscript. All authors had full access to all the data used in this study and had final responsibility for the decision to submit for publication. We declare that we have no conflicts of interest.

## Supplementary Appendix

### A. List of Supplementary Figures and Tables

#### Figures & Tables

**Figure A1:**
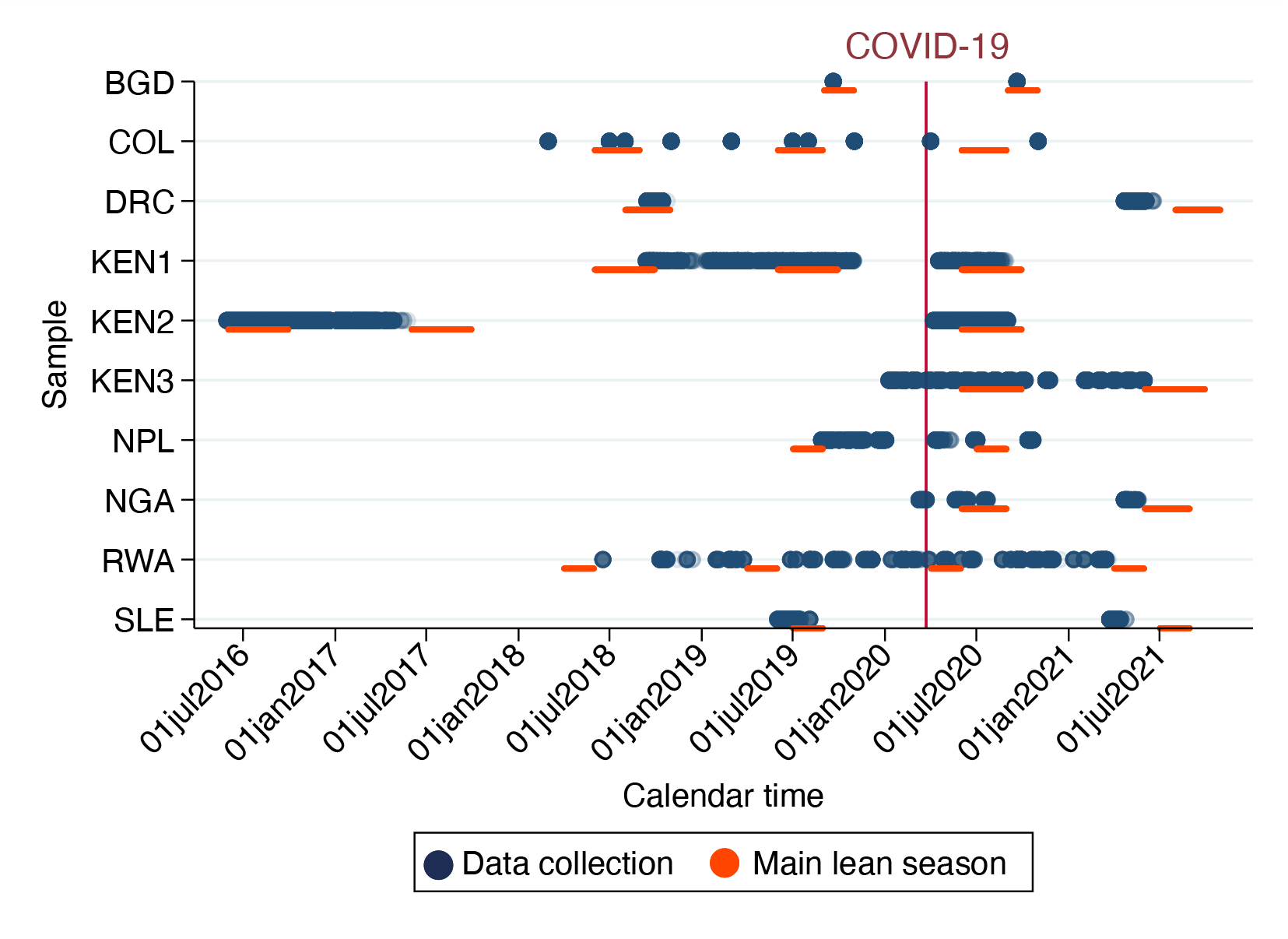
Data collection and lean season timeline. Notes. Figure plots the data collection and lean season timeline across samples to show which crop cycle the COVID-19 arrived in the countries. The Y-axis shows the sample. The X-axis shows the calendar time. Dark blue color refers to individual surveys during the data collection while dark orange color shows the main lean season across countries and years. The red vertical line shows the onset of the COVID-19 pandemic on 11 March 2020.

**Figure A2:**
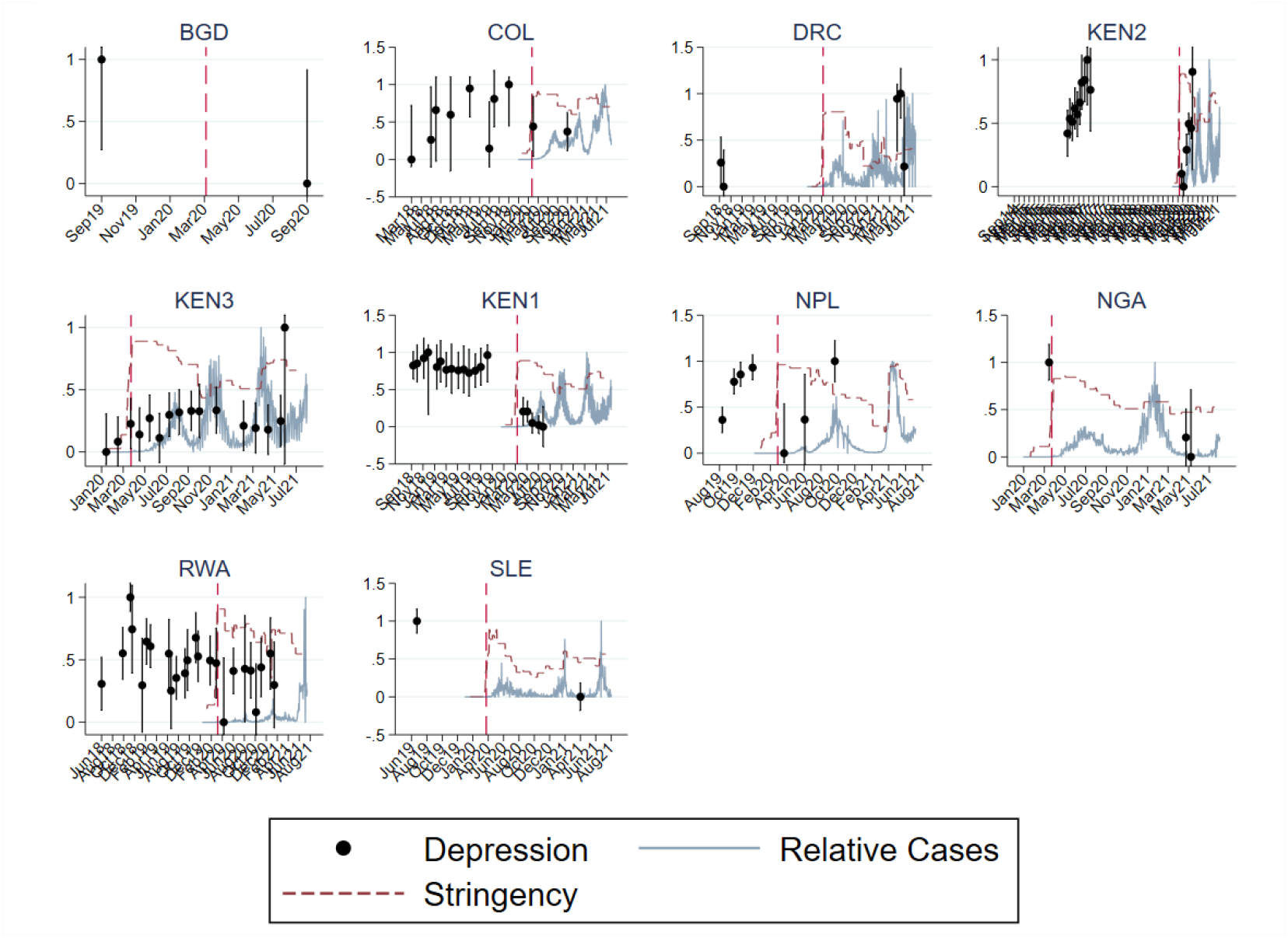
Mental Health, Lockdowns, and Cases.

**Figure A3:**
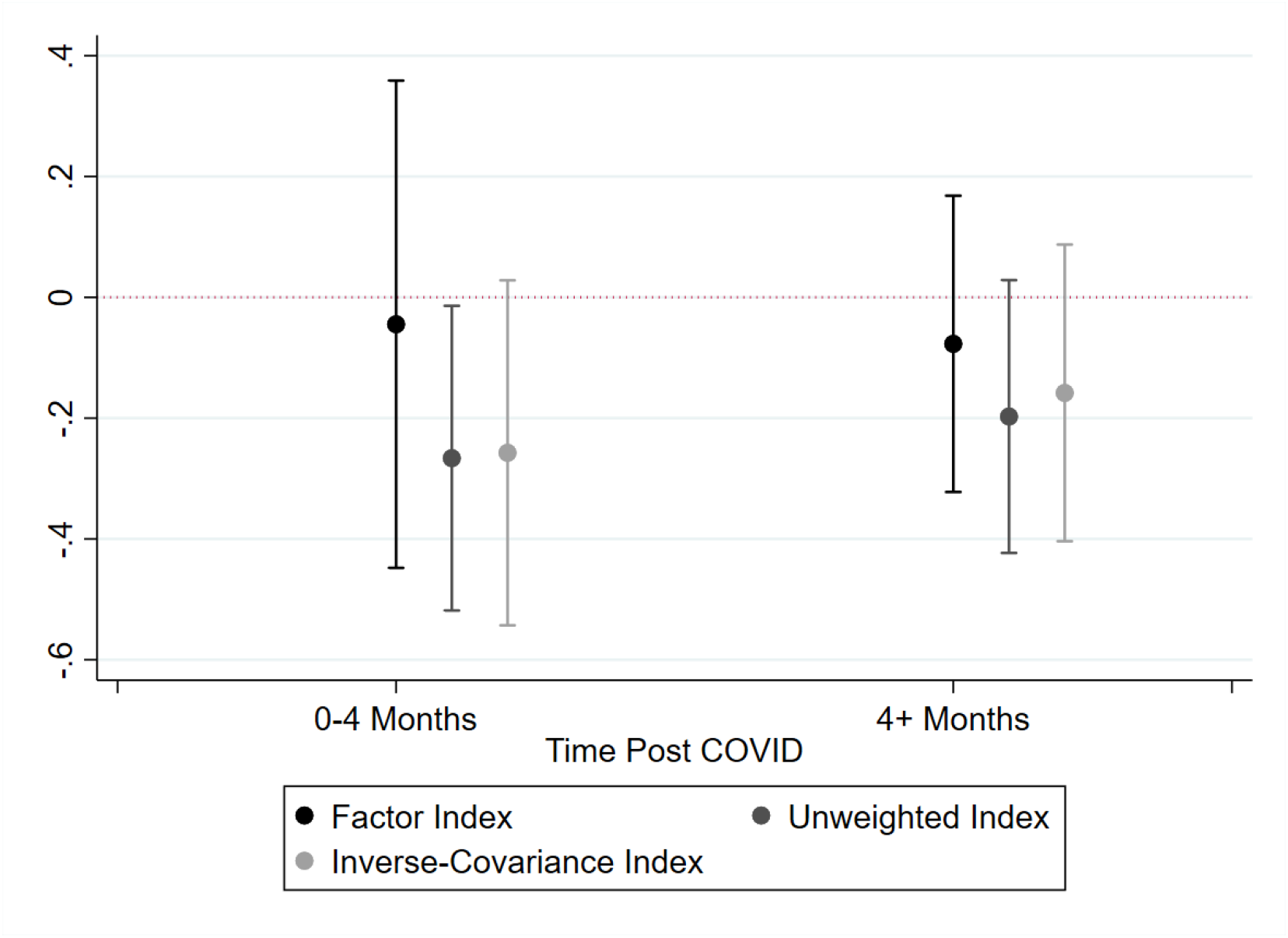
Random Effects Aggregates With Individual Fixed Effects and No Additional Controls. Notes. Figure plots the average change in mental health pre-post COVID across samples using a common estimation strategy that uses individual fixed-effects without additional controls for all samples. The Y-axis is the change in mental health relative to pre-COVID. The X-axis shows time after the onset of COVID discretized into two groups: 0-4 months, and more than 4 months. Point estimates are represented as dots with 95% confidence intervals as whiskers. The color of the points indicates which method of weighting our indices was used: the factor loadings (black, left), the unweighted index (dark gray, middle) or the inverse-covariance weights (light gray, right). The average effect sizes and confidence intervals are estimated from a random-effects model using restricted maximum likelihood to estimate the variance in true effects across samples.

**Table A1:** Description of household survey data samples used in the analysis.

| Country and COVID events | Projects | Households | Survey dates |
| --- | --- | --- | --- |
| **Bangladesh**<br>First case: March 8, 2020<br>Total cases (July 15, 2021): 1,071,774<br>Schools closed: March 17, 2020-September 13, 2021<br>Lockdown: March 26–May 30 | BGD. Rohingya refugees from Myanmar: Refugee camp households in Bangladesh | 3,517 | Sept. 2019 - Oct. 2020 |
| **Colombia**<br>First case: March 6, 2020<br>Total cases (July 15, 2021): 4,583,442<br>Schools closed: March 24, 2020<br>Lockdown: March 24, 2020–September 1, 2020 | COL. Forcibly displaced primary caregivers in psychosocial support program: Caregivers of young children (ages 3-5) enrolled in public Early Childhood Development Centers in Tumaco, Colombia | 662 | March 2018 - Nov. 2020 |
| **Democratic Republic of the Congo**<br>First case: March 6, 2020<br>Total cases (July 15, 2021): 45,211<br>Schools closed: March 28, 2020 - August 15, 2020<br>Lockdown: March 28, 2020 - August 15, 2020 | DRC. Randomly selected Respondents from 80 villages in Nyangezi, Katana, and Walikale Health Zones in Eastern DRC | 1,414 | 14 Sept. 2018 - 19 June 2021 |
| **Kenya**<br>First case: March 13, 2020<br>Total cases (July 15, 2021): 191,020<br>Schools closed: March 20, 2020 - January 4, 2021<br>Curfew: March 27, 2020 - October 20, 2021 | KEN 1. Participants in child health and human capital interventions: 6,500 Kenyans who were received one or more randomized health, skills training, and financial capital interventions during childhood and adolescence | 2,677 | 12 Sep 2018 - 22 Feb 2020; 17 Apr 2020 - 2 Sep 2020 |
|  | KEN 2. Rural Households in NGO Cash Transfer Study: 653 rural villages taking part in an unconditional cash transfer program in Siaya County | 7,585 | 30 May 2016 - 25 May 2017; 07 April 2020 - 02 Sept. 2020 |
|  | KEN 3. Rural households in Financial and Health Diaries study (i-PUSH): Households with pregnant women or children under 4 years old in Kakamega and Kisumu County | 574 | 01 Oct. 2019 - 15 July 2021 |
| **Nepal**<br>First case: January 23, 2020<br>Total cases (July 15, 2021): 662,570<br>Schools closed: March 19, 2020-September 17, 2021<br>Lockdown: March 24, 2020-July 21, 2020 | NPL. Agricultural Households in Western Terai: Rural households in the districts of Kailali and Kanchanpur | 1,372 | 27 Aug. 2019 - 22 Oct. 2020 |
| **Nigeria**<br>First case: February 27, 2020<br>Total cases (July 15, 2021): 169,074<br>Schools closed: March 26, 2020 - September 5, 2021<br>Partial lockdowns: March 29, 2020 - May 18, 2020 | NGA. Irregular Migration and Misinformation in Nigeria: Young adults in Edo and Delta states | 217 | 09 March 2020 - 21 May 2021 |
| **Rwanda**<br>First case: March 14, 2020<br>Total cases (July 15, 2021): 51,625<br>Schools closed: March 16, 2020 - February, 2021<br>Lockdown: March 21, 2020 - May 4, 2020 | RWA. Participants in 100WEEKS cash transfer program: Women in the lowest quantile of income in Musanze, Rwanda. | 336 | 25 June 2016 - 01 April 2021 |
| **Sierra Leone**<br>First case: March 31, 2020<br>Total cases (July 15, 2021): 6,122<br>Schools closed: March 31, 2020 - October 5, 2020<br>Lockdown: April 5–7, 2020, May 3–5, 2020 | SLE. Participants in the Sierra Leone Rural Electrification project that installs solar mini-grids | 2,808 | 01 June 2019 - 25 April 2021 |

**Table A2:** Question wording and answer options for depression measurement.

| Study | Question | Validated screening tool | Reference period |
| --- | --- | --- | --- |
| BGD | • How often did you feel so sad that nothing could cheer you up?<br>• I could not shake off the blues.<br>• I felt depressed.<br>• I felt sad<br>• I could not get going<br>• Nothing made me happy | CES-D-20, Cronbach's $\alpha$ (Scale reliability coefficient)= 0.948 | Last week |
| COL | • Loss of sexual interest or pleasure<br>• Feeling low in energy or slowed down<br>• Thoughts of ending your life<br>• Crying easily<br>• Feeling of being trapped or caught<br>• Blaming yourself for things<br>• Feeling lonely<br>• Feeling blue<br>• Worrying too much about things<br>• Feeling no interest in things<br>• Feeling hopeless about the future<br>• Feeling everything is an effort<br>• Feelings of worthlessness | SCL-90, Cronbach's $\alpha$ =0.855 | Last 4 weeks |
| DRC | • Have you been bothered by these problems?<br>• Feeling low in energy or slowed down.<br>• Blaming yourself for things<br>• Crying easily<br>• Feeling sad or blue<br>• Feeling no interest in things/less interest in daily activities | HSCL-25, Cronbach's $\alpha$ = 0.943 | Last 4 weeks |
| KEN1 | • In past 7 days, how often have you felt depressed? | Subquestion from CESD-10 | Last week |
| KEN2 | • In the past week, I felt depressed and troubled in my mind.<br>• In the past week, I felt lonely.<br>• In the past 7 days, how often have you felt hopeful about the future? | Subquestions from CESD-10 | Last week |
| KEN3 | • Did you sleep well?<br>• Were you happy?<br>• Did you have trouble concentrating?<br>• Do you feel hopeful about the future?<br>• Did you feel that everything you did was an effort?<br>• Did you feel lonely?<br>• Did you feel depressed/stressed?<br>• Did you feel that you could not 'get going'?<br>• Were you bothered by things that don't usually bother you?<br>• Did you feel fearful? | CES-D-10, Cronbach's $\alpha$ = 0.773 | Last week |
| NPL | • In the past 14 days how often have you felt sad or depressed?<br>• In the past 14 days, how often did you have difficulty sleeping at night?<br>• In the past 14 days how often have you felt stressed or worried? | N/A | Last 2 weeks |
| NGA | • I have felt cheerful and in good spirits<br>• I have felt calm and relaxed<br>• I have felt active and vigorous<br>• I woke up feeling fresh and rested<br>• My daily life has been filled with things that interest me | WHO-5 Well-being Index, Cronbach's $\alpha$ = 0.82 | Last 2 weeks |
| RWA | • In the past 7 days, were you/did you<br>• ...lost a lot of sleep because of worries?<br>• ...felt that you couldn't overcome your difficulties?<br>• ...able to face up to your problems? | N/A | Last week |
| SLE | • Feeling happy<br>• Feeling satisfied<br>• Facing difficulties/ feeling a lack of control over life | N/A | N/A |

Notes. This table presents question wording and answer options for mental health variables used in analysis. In BGD, COL, DRC, KEN3 and NGA samples, we followed the prescribed calculation method of each scale. In the remaining samples we constructed indices of the available depression items as explained in the Data and Methods section. In BGD and KEN3, the total CES-D score (range: 0-80 and 0-30, respectively) is calculated by summing all the items. In COL, the SCL-90 score is calculated by dividing the raw score to number of questions in the depression domain (n=13). In DRC, the final score is calculated by dividing the sum of the scores of all the items by 25 (the final score ranges from 1.00 to 4.00), where at least 22 items had to be answered for the assessment to be considered valid. In NGA, the total raw score, ranging from 0 to 25, is multiplied by 4 to give the final score, with 0 representing worst possible, whereas a score of 100 representing best possible quality of life. All scores are reverse coded and standardized by mean=0, standard deviation=1. The higher scores represents better mental well-being for each scale.

**Table A3:** Best Estimates from Each Sample (ICW Index)

|  | Season + Time Trend |  |  | Seasonal Food Security Ctrl |  | Time Control | Pre-Post Only |  |  |  |
| --- | --- | --- | --- | --- | --- | --- | --- | --- | --- | --- |
|  | (1) | (2) | (3) | (4) | (5) | (6) | (7) | (8) | (9) | (10) |
|  | RWA | COL | KEN1 | KEN2 | NPL | KEN3 | BGD | NGA | SLE | DRC |
| 0 |  |  |  |  |  |  |  |  |  |  |
| 0-2 months |  | -0.201**<br>(0.0907) | -0.705***<br>(0.193) | -0.304***<br>(0.0156) | -0.189***<br>(0.0696) | -0.0190<br>(0.0478) |  |  |  |  |
| 2-4 months | -0.306<br>(0.624) |  | -0.873***<br>(0.183) | -0.271***<br>(0.0268) | -0.0598<br>(0.0640) | -0.152*<br>(0.0754) |  |  |  |  |
| 4-6 months | -0.614<br>(0.456) |  | -0.851***<br>(0.208) | -0.275***<br>(0.0400) |  |  | 0.0828**<br>(0.0381) |  |  |  |
| 6-9 months | -0.384<br>(0.388) | -0.336***<br>(0.101) |  |  | 0.0594*<br>(0.0343) |  |  |  |  |  |
| 9-12 months | -0.558<br>(0.477) |  |  |  |  |  |  |  |  |  |
| 12-15 months |  |  |  |  |  |  |  | -0.351***<br>(0.0828) | 0.100**<br>(0.0389) | 0.275***<br>(0.0482) |
| Year | 0.492***<br>(0.189) | 0.147**<br>(0.0599) | 0.0172<br>(0.161) |  |  | 0.878***<br>(0.153) |  |  |  |  |
| Seasonal Food Security |  |  |  | 0.423***<br>(0.105) | 0.137***<br>(0.0229) |  |  |  |  |  |
| Observations | 1532 | 2503 | 5405 | 24899 | 13143 | 8342 | 6311 | 1081 | 5978 | 3183 |

Standard errors in parentheses
\* $p < .1$ , \*\* $p < .05$ , \*\*\* $p < .01$

**Table A4:**
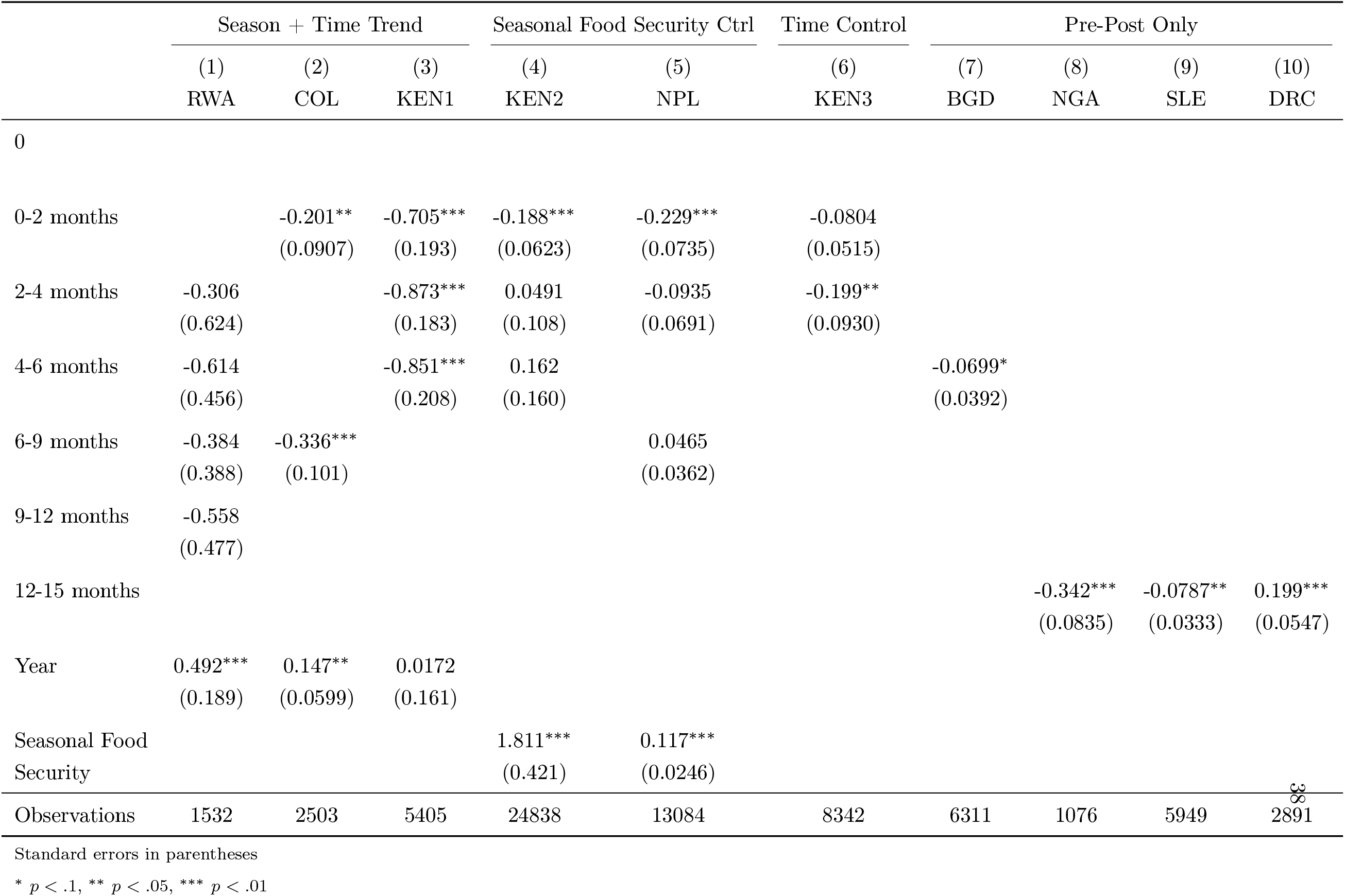
Best Estimates from Each Sample (Factor Index)

**Table A5:** Pre-post differences in depression index (Factor index)

|  | (1)<br>BGD | (2)<br>COL | (3)<br>KEN2 | (4)<br>KEN3 | (5)<br>KEN1 | (6)<br>NPL | (7)<br>NGA | (8)<br>RWA | (9)<br>SLE | (10)<br>DRC |
| --- | --- | --- | --- | --- | --- | --- | --- | --- | --- | --- |
| 0 |  |  |  |  |  |  |  |  |  |  |
| 0-2 months |  | -0.0499<br>(0.0572) | -0.0255<br>(0.0239) | 0.0489<br>(0.0410) | -0.731***<br>(0.0776) | -0.244***<br>(0.0838) |  |  |  |  |
| 2-4 months |  |  | 0.183***<br>(0.0286) | 0.0467<br>(0.0442) | -0.922***<br>(0.0672) | -0.131*<br>(0.0789) |  | 0.818***<br>(0.200) |  |  |
| 4-6 months | -0.0699*<br>(0.0392) |  | 0.351***<br>(0.0286) | 0.0850**<br>(0.0425) | -0.934***<br>(0.101) |  |  | 0.300<br>(0.337) |  |  |
| 6-9 months |  | -0.0676<br>(0.0452) |  | 0.0958**<br>(0.0458) |  | 0.0615<br>(0.0411) |  | 0.231<br>(0.236) |  |  |
| 9-12 months |  |  |  | 0.0377<br>(0.0493) |  |  |  | -0.0240<br>(0.270) |  |  |
| 12-15 months |  |  |  | 0.0558<br>(0.0489) |  |  | -0.342***<br>(0.0835) |  | -0.0787**<br>(0.0333) | 0.199***<br>(0.0547) |
| Observations | 6311 | 2503 | 24909 | 8342 | 5405 | 13084 | 1076 | 1532 | 5949 | 2891 |

Standard errors in parentheses
\* $p < .1$ , \*\* $p < .05$ , \*\*\* $p < .01$

**Table A6:** Pre-post differences in depression index (ICW index)

|  | (1)<br>BGD | (2)<br>COL | (3)<br>KEN2 | (4)<br>KEN3 | (5)<br>KEN1 | (6)<br>NPL | (7)<br>NGA | (8)<br>RWA | (9)<br>SLE | (10)<br>DRC |
| --- | --- | --- | --- | --- | --- | --- | --- | --- | --- | --- |
| 0 |  |  |  |  |  |  |  |  |  |  |
| 0-2 months |  | -0.00959<br>(0.0556) | -0.455***<br>(0.0227) | 0.158***<br>(0.0437) | -0.731***<br>(0.0776) | -0.205**<br>(0.0832) |  | -0.269<br>(0.352) |  |  |
| 2-4 months |  |  | -0.309***<br>(0.0269) | 0.185***<br>(0.0468) | -0.922***<br>(0.0672) | -0.100<br>(0.0765) |  | -0.323**<br>(0.147) |  |  |
| 4-6 months | 0.0828**<br>(0.0381) |  | -0.212***<br>(0.0276) | 0.231***<br>(0.0447) | -0.934***<br>(0.101) |  |  | -0.644***<br>(0.225) |  |  |
| 6-9 months |  | -0.0173<br>(0.0450) |  | 0.205***<br>(0.0475) |  | 0.0815**<br>(0.0408) |  | -0.386**<br>(0.187) |  |  |
| 9-12 months |  |  |  | 0.120**<br>(0.0522) |  |  |  | -0.774***<br>(0.177) |  |  |
| 12-15 months |  |  |  | 0.166***<br>(0.0512) |  |  | -0.351***<br>(0.0828) |  | 0.100**<br>(0.0389) | 0.275***<br>(0.0482) |
| Observations | 6311 | 2503 | 24970 | 8342 | 5405 | 13143 | 1081 | 1712 | 5978 | 3183 |

Standard errors in parentheses
\* $p < .1$ , \*\* $p < .05$ , \*\*\* $p < .01$

**Table A7:** Pre-post differences in depression index (Unweighted index)

|  | (1)<br>BGD | (2)<br>COL | (3)<br>KEN2 | (4)<br>KEN3 | (5)<br>KEN1 | (6)<br>NPL | (7)<br>NGA | (8)<br>RWA | (9)<br>SLE | (10)<br>DRC |
| --- | --- | --- | --- | --- | --- | --- | --- | --- | --- | --- |
| 0 |  |  |  |  |  |  |  |  |  |  |
| 0-2 months |  | -0.0650<br>(0.0569) | -0.353***<br>(0.0226) | 0.0483<br>(0.0405) | -0.731***<br>(0.0776) | -0.210**<br>(0.0832) |  | -0.257<br>(0.357) |  |  |
| 2-4 months |  |  | -0.179***<br>(0.0268) | 0.0461<br>(0.0436) | -0.922***<br>(0.0672) | -0.105<br>(0.0766) |  | -0.299**<br>(0.149) |  |  |
| 4-6 months | -0.0466<br>(0.0391) |  | -0.0737***<br>(0.0275) | 0.0839**<br>(0.0420) | -0.934***<br>(0.101) |  |  | -0.624***<br>(0.226) |  |  |
| 6-9 months |  | -0.0672<br>(0.0451) |  | 0.0946**<br>(0.0452) |  | 0.0790*<br>(0.0408) |  | -0.369*<br>(0.189) |  |  |
| 9-12 months |  |  |  | 0.0372<br>(0.0486) |  |  |  | -0.761***<br>(0.179) |  |  |
| 12-15 months |  |  |  | 0.0551<br>(0.0483) |  |  | -0.355***<br>(0.0830) |  | -0.199***<br>(0.0344) | 0.242***<br>(0.0479) |
| Observations | 6311 | 2503 | 24970 | 8342 | 5405 | 13143 | 1081 | 1712 | 6036 | 3133 |

Standard errors in parentheses
\* $p < .1$ , \*\* $p < .05$ , \*\*\* $p < .01$

### B. Sample Descriptions and COVID-19 Experiences

In this Appendix section we describe the economic context, COVID-19-related developments, and sample construction for each sample in the study.

#### B.1 Bangladesh

##### COVID-19 Experience

*Case History:*

- March 8, 2020: First confirmed cases reported by the Institute of Epidemiology, Disease Control, and Research (IEDCR)
- Total cases: 1,071,774 as of July 15, 2021 (Our World in Data)
- Total deaths: 17,278 as of July 15, 2021 (Our World in Data)

*Mobility Restrictions:*

- March 14, 2020: On-arrival visas suspended for all countries. Ban on flights from all European countries except the United Kingdom
- March 26-May 30, 2020: The Government of Bangladesh (GoB) declared a “national holiday”, limited the availability of public transport, and ordered all public and private offices to remain closed. Only food markets, pharmacies, hospitals, and emergency services were allowed to remain open.
- April 9, 2020: The GoB imposes a “complete lockdown” on Cox’s Bazar District. No entry and exit from the district is permitted.
- June 1, 2020: The GoB divides the country into zones (high risk, moderate, and low risk), based on the number of COVID-19 cases. Movement across areas was restricted.

*Social Distancing:*

- May 31, 2020: Face masks mandatory when outside the home.
- June 27, 2020: Public gatherings prohibited in red zones. Stay at home required for red zones.

*School Closures:*

- March 17, 2020-September 13, 2021: Closure of all schools and universities.

*Social Protection Responses:* Bangladesh launched two major cash transfer programs in response to the pandemic. The assistance program for garment sector workers offered digital payments to 4 million employees at textile factories (Gentilini et al. 2020, Chowdury 2020). Another cash transfer program offered top-up payments to 5 million households who were already receiving government benefits (“PM to launch disbursement of cash aid” 2020). These two programs benefitted approximately 15% of the population (Gentilini et al. 2020).

- March 26: Prime Minister announced an USD 588 million package for export-oriented industries, to be spent on employee salaries
- April 5: Prime Minister announced an USD 8 billion stimulus package for hard-hit industries, small and medium enterprises (SMEs), and emergency incentives for export oriented industries. The GoB announced an expansion of the social safety net programs—including the Vulnerable Group Feeding(VGF) and Vulnerable Group Development (VGD) programs—and reductions in rice prices.

##### Economic Context

*Seasonality and Food security* The Bangladesh survey was conducted in September 2019 and October 2020, which partially overlaps with the lean period. Despite an above-average 2019 harvest, prices of rice during in Bangladesh survey remained well above their prior year levels, linked to pandemic-related increases in demand and concerns about the upcoming 2020 harvest (Global Information and Early Warning System on Food and Agriculture Country Brief: Bangladesh, *Food and Agricultural Organisation of the United Nations*, 10-June-2020).

The Bangladesh post-COVID surveys in this article were conducted after to Tropical Cyclone Amphan, which struck southwestern parts of Bangladesh on 20 May 2020 and caused loss of life and substantial devastation, including livestock and crops.

*Refugees* The over 900,000 Rohingya refugees living primarily in Cox’s Bazar district since 2017 have experienced food insecurity and required humanitarian assistance to meet daily needs since well prior to the Covid-19 pandemic (Global Information and Early Warning System on Food and Agriculture Country Brief: Bangladesh, *Food and Agricultural Organisation of the United Nations*, 10-June-2020).

*Social Protection* The social protection policy environment in Bangladesh is fragmented, with over 114 separate programs providing cash and food transfers to the vulnerable (ILO 2020).

##### B.1.1 Bangladesh BGD, Rohingya refugees from Myanmar: Refugee camp households in Bangladesh

**Project Title:** Mental wellbeing of displaced Rohingya women: a cluster randomised controlled trial (RCT)

**Target Population**: Rohingya women with a child aged 0-2 years.

**Original Study Design:** Cluster RCT.

*Sampling Frame:* Geographic-level information, which is blocks within the camps, was considered for randomisation. At the time of randomization, there were about 2000 blocks. The number of blocks required in each arm was computed with an improvement in mental health by 0.05 units (on a mental health index scale between 0 and 1) at 5% significance level with 80% power. A minimum of 112 blocks with 1,568 mother-child pairs are required in each arm to meet this requirement. A total of 251 blocks were selected, of which 137 were assigned to the treatment (54%) and 114 were assigned to the control group (46%). Treatment and control blocks have been selected in a manner that they were not too close to each other, minimizing potential contamination.

**COVID-19 Survey Design:** Phone surveys.

*Sampling Frame:* At baseline, a total of 3499 participants were surveyed: 1,911 from the treatment and 1,588 from the control groups. At endline, 2,845 participants were surveyed, 1,679 from the treatment and 1,166 from the control arm.

*Survey Dates:* Baseline in September 2019; endline in October 2020.

*Sample size, tracking and attrition:* About 19% participants attired, mostly due to lack of access to mobile phones.

*Median survey time:* Median survey time is unknown as survey was conducted over the phone and there were other components on child health.

*Sampling Weights:* N/A.

*IRB Approval:* This research was approved by BRAC University (ref no. 2019-028-ER).

#### B.2 Colombia

##### COVID-19 Experience

*Case History:*

- March 6, 2020: First confirmed case
- Total cases: 4,583,442 as of July 15, 2021 (Our World in Data)
- Total deaths: 114,833 as of July 15, 2021 (Our World in Data)

*Mobility Restrictions:*

- March 17-July 1, 2020: Borders closed.
- March 24, 2020: Mandatory preventive isolation implemented throughout the country, which allows one member of the family to leave to buy food, medicine and carry out financial transactions (extended to September 1).
- March 24, 2020: General lockdown until April 13 (extended to September 1).
- May 6, 2020: Three exceptions for internal movement: i) humanitarian emergency; ii) transport of cargo or merchandise; iii) fortuitous event or force majeure.

*School Closures:*

- March 24, 2020: Schools are closed.

*Social Distancing:*

- March 16, 2020: Public events for more than 50 people are banned.
- March 24, 2020: Mandatory preventive isolation implemented throughout the country. Social events and activities are prohibited including religious services involving crowds or gatherings, group sports, gyms, bars and discos, and cinemas and theaters. Restaurants can only provide take-away orders. Consumption of alcohol in open spaces is banned.
- April 4, 2020: The Government mandates the use of face masks in public transit and in areas of high volume such as supermarkets, banks and pharmacies. Face masks are mandatory for people with respiratory symptoms and vulnerable groups such as adults over 70.

*Social Protection Responses:*

- March 25, 2020: Individuals in strata 1 and 2 can defer payment of energy and gas bills up to 36 months during mandatory preventive isolation. Cash transfers and food aid provided to households, youth and the elderly.
- In March 2020, Colombia rolled out a new unconditional cash transfer program, *Devolución del IVA*, benefitting one million low-income households. The transfer is paid every two months to recipients of *Familias en Acción*, and *Colombia Mayor*. The 75,000 peso (USD 20) additional transfer represents 8% of the monthly minimum wage. Approximately 27% of the population has received this cash transfer program (Gentilini et al. 2020).

##### Economic Context

*Seasonality and Food Security* The Colombia post-COVID survey was conducted during the beginning of a lean period. The price of rice reached a record high in April 2020, shortly after COVID-19 hit the region. Prices remained 40% above year-earlier values by June 2020.

*Conflict* The Coordination Platform for Refugees and Migrants for Venezuela estimated that nearly 5 million people have fled from the country as of mid-March 2020. Colombia is the main host country of refugees and migrants from Venezuela (Bolivarian Republic of Venezuela), with an estimated population of nearly 1.8 million. According to a February 2020 World Food Program survey, more than 20 percent of the migrant population in the four departments that border with Venezuela were severely food insecure. Migrant households have minimal access to employment, increasing food insecurity. On top of the inflow of Venezuelan refugees, Colombia has 8.2 million Internally Displaced Persons, which accounts to 16% of the population and is the highest number of IDP worldwide. Despite a generous and progressive framework that started in 1997, IDP are still more vulnerable than the urban and rural poor. In 2014, poverty and extreme poverty rates were 1.7 and 3.5 times higher for IDP relative to national averages. Conflict and force displacement are still happening and during the pandemic (between 2020 and 2021), internal forced displacement rose by 181%.

*Social Protection* Colombia provides several social protection programs, including a conditional cash transfer program, *Familias en Acción*, and *Colombia Mayor*, a non-contributory pension program for low-income senior citizens.

##### B.2.1 Colombia COL, Primary caregivers in Tumaco, Colombia

**Project Title:** Semillas de Apego: a psychosocial group model to promote maternal mental health and early childhood development

**Target Population**: Primary caregivers of young children (ages 3-5) enrolled in public Early Childhood Development Centers in Tumaco, Colombia. Caregivers had a prior exposure to violence and forced displacement.

**Original Study Design:** Original study is based on cluster-based randomization to the ‘Semillas de Apego’ psychosocial support program. ECDCs were first randomised to the treatment or control groups. Second, all caregivers of children who were attending an ECDC were randomly allocated to four sequential cohorts following a phased-in approach. All caregivers received regular child and family services provided by the ECDC, while caregivers in the treatment group received an invitation to participate in the program. In every cohort, participants completed assessments at baseline, and 1 and 8-month follow-ups after the program had been completed with the treatment group. Baseline survey included a detailed socioeconomic module, questionnaire on exposure to violence and forced displacement, and scales on child-parent interactions and early-childhood development. Follow-up surveys included the latter scales and short modules to record changes in socioeconomic characteristics and exposure to violence between waves.

*Sampling Frame:* Universe of caregivers whose children were served by 18 public ECDC in Tumaco, Nariño. These 18 ECDC serve 80% of the 1600 families in the municipality whose children are enrolled across public ECDCs. Sampling in each cohort was designed using administrative-level data provided by each ECDC. 1376 primary caregivers participated over the course of the programme in the treatment (n=714) or control (n=662) groups.

**COVID-19 Survey Design:** Phone surveys with selected items from SCL-90-R battery administered in in-person data collection, Parenting Stress Index short form questionnaire, and a module on Covid-related stressors.

*Sampling Frame:* Sample of 1376 caregivers who had participated in the prior (pre-pandemic) RCT and data collection.

*Survey Dates:* Baseline data was collected in March, 2018 (Cohort 1); July, 2018 (Cohort 2); March, 2019 (Cohort 3); and July, 2019 (Cohort 4). 1-month follow-up data was collected in August, 2018 (Cohort 1); November, 2018 (Cohort 2); August, 2019 (Cohort 3); and November, 2019 (Cohort 4). 8-month follow-up data was collected in Marc, 2019 (Cohort 1); and August, 2018 (Cohort 2) and was scheduled to be collected in March, 2020 (Cohort 3); and August, 2020 (Cohort 4) but field work was interrupted because of the pandemic and associated lockdowns. Two short phone surveys were then administered to assess the mental health impact of the pandemic. First phone survey was administered between April 7 and 21, 2020 (Cohorts 3 and 4) and the second one between November 12 - December 12, 2020 (All cohorts).

*Sample size, tracking and attrition:* 1,376 caregivers. 90% (1245 of 1376) participants completed the 8-month follow-up survey (in-person or April phone survey). Attrition was not different between the in person (9%) and phone survey (10%) and was not predicted by baseline characteristics. 7% (1285 of 1376) participants completed the November 2020 phone survey.

*Median survey time:* In person surveys (including child-development battery): 2.5 hours. Phone surveys: 20 minutes.

*Sampling Weights:* N/A.

*IRB Approval:* Original study design approved by the Ethics Committee of the Universidad de los Andes, Colombia (protocol 786, 2017). Inclusion of Covid-19 surveys approved under the same study protocol in April 2020.

#### B.3 Democratic Republic of Congo

##### COVID-19 Experience

*Case History:*

- March 14, 2020: First confirmed case
- Total cases: 45,211 as of July 15, 2021 (Our World in Data)
- Total deaths: 984 as of July 15, 2021 (Our World in Data)

*Mobility Restrictions:*

- March 21, 2020: All borders are closed and international passenger flights are suspended (U.S. Embassy in the Rebublic of the Congo, 2020).
- March 28, 2020: Places of worship, bars and nightclubs are closed.
- March 28, 2020: Workplaces closing except for workers providing essential goods and services.
- March 30, 2020: Lockdown measures include suspension of non-essential travel and a curfew until May 31, 2020 (FCDO, 2020).
- May 18, 2020: Nationwide overnight curfew between 8pm and 5am, revised to 10pm to 5am in July 28, 2020, ended in August 15, 2020.

*School Closures:*

- March 28, 2020: Schools and higher education institutions are closed.
- May 18, 2020: Schools are reopened for primary schools’ final year class and graduation class (IMF, 2020).
- June 1, 2020: The school grades of CM2, 3ieme and Terminale are repopened.
- August 10, 2020: All schools are reopened (Anadolu Agency, 2020).

*Social Distancing:*

- March 28, 2020: Gatherings of more than 50 people are prohibited.
- August 15, 2020: Places of worship were allowed to reopen (U.S. Embassy in the Rebublic of the Congo, 2020).

*Social Protection Responses:*

- March 31, 2020: Free water and electricity for all households during the confinement period. Financial aid of 4 billion granted to households and people experiencing poverty. Government is replacing less than 50% of lost salary.

##### Economic Context

*Seasonality and Food Security* The DRC post-COVID surveys were conducted between April through June 2021, during the lean season. Crop output in 2020 was lower than typical owing to floods, pests, violence, and COVID-19 preventative efforts (FAO, 2020). Between April and July 2020, the prices of imported food goods rose dramatically. Decrease in cash crop exports as a consequence of poor demand from importing countries and a slowdown in trade flows as a result of COVID-19 resulted in a fall in foreign exchange revenues and prices to rise.

*Conflict* Weak governance and the prevalence of many armed groups have subjected Congolese civilians to widespread rape and sexual violence, massive human rights violations, and extreme poverty. Millions of civilians have been forced to flee the fighting: the United Nations estimates there are currently 4.5 million internally displaced persons in the DRC.

*Social Protection* In 2017, a strategic framework was established by the government to adopt a comprehensive national social protection policy. However, the programs remained fragmented, poorly funded, and failed to adequately meet the needs of the poorest and most vulnerable in a context of deep fragility. As of 2018, the poverty rate of the country was 63.4%.

##### B.3.1 Democratic Republic of Congo (DRC), Impact evaluation of Community Based Trauma Healing

**Project Title:** Impact evaluation of Community Based Trauma Healing in the Democratic Republic of Congo

**Target Population**: Households in 160 communities in Nyangezi, Katana and Walikale Health Zones in Eastern DRC

**Original Study Design:** To evaluate the impact of a Community Based Trauma Healing (CBTH) implemented in Eastern DRC, as part of a wider program that aims to reduce Gender Based Violence. CBTH aims to address communal trauma through community engagement, building social cohesion and exploring and deconstructing negative stereotypes. The program recruited local Community Trauma Healers that were trained and supported by an international NGO to organize a community sessions and celebrations. During the session, the program aimed to help community members heal from post-conflict trauma resulting from, among other things, death, injury, (systematic) rape, indigenous conflict, internal displacement and inheritance conflicts. These community “detraumatisme” sessions brought together (potential) victims, perpetrators, traditional leaders and faith-based leaders. CBTH teaches community members strategies to control emotions, and reduce risks of GBV. We use a cluster randomized trial to evaluate the impact of CBTH in 80 randomly selected communities in Eastern DRC.

*Sampling Frame:* To select the villages, first, the implementing partner selected three health zones, and then created a list of 40 health areas that met three criteria: they were receiving wider program interventions, they were relatively accessible, and had not anticipated major security risks that might undermine or interrupt program and evaluation. Within this subset of health areas, 160 eligible villages were identified on the basis of their accessibility, relative security, population size and household count, and a limited presence of other projects that might contaminate any impact of the intervention. In each health area, the implementing partner identified four villages. Pre-COVID (baseline) data was collected during September and October 2018 each of the 160 villages. Within each village, field teams used a “random-walk” methodology to randomly select households. Enumerators interviewed every 3rd household on the right in rural areas and every 5th household on the right in semi-urban areas, starting from a predetermined village landmark (market, administrative building, well, tree, etc.) Prior to the start of daily data collection, the team supervisor was in charge of dispersing enumerators around the village so that they could cover all the area and that the sample would not be confined to a small portion of the observation area. In each selected household, enumerators interviewed a random adult respondent (age 18 or over) selected from within the household (using a kish-grid method programmed on the tablet). As per the programming, and to ensure the highest level of responsiveness on sensitive topics, female enumerators only interviewed female respondents and male enumerators only interviewed male respondents. In total, we collected baseline data from 3678 respondents.

**COVID-19 Survey Design:** Post COVID (endline) data was collected during April through June 2021. We aimed to resurvey all baseline respondents. When enumerators were unable to interview a baseline respondent, they recorded the reason (unable to locate respondent, respondent temporarily away, permanently away, died, or refusal). If the enumerator was able to locate the household but not the specific baseline respondent in that household, then the enumerator attempted to find another eligible respondent (18 or older) of the same sex as the baseline respondent within the household that was closest in age. Failing that, the enumerator would replace the household with the household to the right of the original household and would identify.

*Sampling Frame:* This study uses data from the control group only, comprising 80 villages, comprising 1414 respondents.

*Survey Dates:* September and October 2018 and April through June 2021.

*Sample Size, Tracking and Attrition:* Pre-COVID data was collected under 1771 respondents. Followup data was collected under 1771 respondents.

*Median Survey Time:* 67 minutes

*Sampling Weights:* None.

*IRB Approval:* The research protocol for this study was approved by the NORC Institutional Review Board (IRB00000967), Project Number 7554.059.01, IRB Protocol Number 18.08.13 on 09/10/2018.

#### B.4 Kenya

##### COVID-19 Experience

*Case History:*

- First confirmed case: March 13, 2020
- Total cases: 191,020 as of July 15, 2021 (Our World in Data)
- Total deaths: 3,746 as of July 15, 2021 (Our World in Data)

*Mobility Restrictions:*

- March 15, 2020: Entry restricted to citizens and foreigners with valid residence permits, who must self-quarantine upon arrival. Non-essential government and businesses directed to begin working from home.
- March 25, 2020: National and international flights halted.
- March 27, 2020: Start of dusk to dawn curfew (7pm-5am). Schedule changes to 9pm-4am on June 7; to 11pm–4am on September 28, and to 10pm-4am on November 4. Extended to October 20, 2021.
- April 6, 2020: Partial mobility restrictions begin; movement in and out of Nairobi Metropolitan Area restricted. On April 8, 2020, Mombasa, Kilifi and Kwale were was added to this list. Mandera was added on April 22, 2020. Restrictions were lifted for Kilifi and Kwale on June 6, 2020; they remain in place for Nairobi, Mombasa and Mandera.
- In addition, from May 6, 2020 - June 6, 2020, movement into and out of the Eastleigh in Nairobi and Old Town in Mombasa was restricted.

*Social Distancing:*

- March 15, 2020: Limits on large gatherings, church attendance, closure of courts; Safaricom suspends mobile money transaction fees to encourage cashless transactions.
- April 12, 2020: Mask wearing in public spaces mandatory.
- July 6, 2020: Re-opening for congregational worship and public (in-person) worship.

*School Closures:*

- March 20, 2020 - January 4, 2021: Closure of all schools and higher education institutions.

*Social Protection Responses:*

- March 25, 2020: Announcement of tax breaks for households and enterprises, accelerated payments of bills and tax credits, additional appropriations for cash transfers to the elderly, orphans and vulnerable households by the Ministry of Labour and Social Protection.
- May 23, 2020: Stimulus package announced, including spending on infrastructure, tourism, education sector and SMEs.
- There are reports that beneficiaries of the old age pension (Inua Jamii) program have received additional payments during the pandemic (Gentilini et al. 2020), but other sources suggest that they only received their standard payments (Chebii & Oyunge 2020).
- A public works program temporarily employed 26,000 young people to clean and sanitize poor urban neighborhoods (Matiang’i 2020).

##### Economic Context

*Seasonality and Food Security* The three Kenya post-COVID surveys were conducted as the country entered the lean season, following a below average cereal crop harvest in 2019. A severe desert locust outbreak, the worst in 70 years, has affected large areas in the north and central pastoral areas of the country (KEN3), but households in two Kenya samples (KEN1 and KEN2) are mostly unaffected (Global Information and Early Warning System on Food and Agriculture Country Brief: Kenya, *Food and Agricultural Organisation of the United Nations*, 12-May-2020).

*Social Protection* Kenya has four major cash transfer programs for people who are disabled, elderly, vulnerable children, or at risk of seasonal hunger, coordinated under the National Safety Net Program (National Social Protection Secretariat, n.d.). Approximately 5.7% of the population benefits from these programs (Beegle, Coudousel & Monsalve 2018).

##### B.4.1 Kenya KEN1, Participants in child health and human capital interventions in Kenya

**Project Title:** Kenya Life Panel Survey (KLPS)

**Target Population**: KLPS is a longitudinal dataset of over 6,500 Kenyans that participated in randomized child health and human capital interventions and have since been followed into adulthood. Respondents attended primary school in Busia, Kenya; migrants continue to be tracked and the study sample now resides throughout Kenya, Uganda, and elsewhere. Data used here come from the fourth round (KLPS-4), over 20 years after the first intervention, and phone surveys conducted during the COVID-19 pandemic.

**Original Study Design:** The KLPS sample comprises of individuals who participated in one of two previous randomized NGO programs: the Primary School Deworming Program (PSDP), which provided deworming medication to primary school students during 1998–2003 (Miguel and Kremer 2004), and the Girls’ Scholarship Program (GSP), which provided merit scholarships to upper primary school girls in 2001 and 2002 (Kremer, Miguel and Thornton 2009). Approximately 20% subset of these individuals also participated in the vocational training and cash grants programs (known as the Start-up Capital for Youth and Vocational Education Program, or SCY/Voced) during 2009-2014. Data used in this paper come from KLPS Round 4 (KLPS-4), which collected information roughly 20 years after the initial deworming treatment.

*Sampling Frame:* The KLPS sample is split into 2 representative waves for data collection; analyses here focus on “Wave 1” respondents, as these individuals were surveyed in-person from October 2018-February 2020 and again as part of COVID-19 phone surveys.

**COVID-19 Survey Design:** All KLPS respondents were targeted for phone surveys. The KLPS sample was split into five representative “batches”, and each batch was surveyed in sequence in order to provide representative data over time. Phone surveys were shortened from in-person surveys, capturing information on household earnings, consumption aggregates, physical and mental health.

*Sampling Frame:* A representative sample of approximately 7,500 PSDP participants (out of 32,000 pupils) were selected for inclusion in KLPS, along with the fourth round of the KLPS (KLPS-4) focuses on the subsets of the KLPS sample who participated in the PSDP or SCY/Voced interventions.

*Survey Dates:* Previous KLPS survey rounds were conducted in 2003-05 (KLPS-1), 2007-09 (KLPS-2), and 2011-14 (KLPS-3). KLPS-4 E+ Module (economic data) were collected from 2017-19, and KLPS-4 Wave 1 I-Module surveys (health and mental health data) were conducted from October 2018 to February 2020. COVID-19 phone surveys were conducted from April to August 2020.

*Sample size, tracking and attrition:* KLPS-4 I Module Wave 1 sample size: 4,076. KLPS-4 I Module Wave 1 effective survey rate among the non-deceased (accounting for intensive tracking): 89.6%. COVID-19 phone survey rate among Wave 1 respondents: 77.5%.

*Median survey time:* In-person surveys: 146 min; Phone surveys: 37 min.

*Sampling Weights:* Observations are weighted to be representative of the original KLPS population.

*IRB Approval:* UC Berkeley and Maseno University.

##### B.4.2 Kenya KEN2, Rural Households in NGO Cash Transfer Study

**Project Title:** General Equillibrium Effects of Cash Transfers (GE), Siaya County

**Target Population**: All households across 653 rural villages in three subcounties taking part in an unconditional cash transfer program reported in Egger et al. (2019).

##### Original Study Design

The NGO GiveDirectly (GD) provides unconditional cash transfers to poor households; its cash transfer program was randomly assigned at the village level. In treatment villages, poor households meeting a basic means test (33 percent of households) were eligible for the program. Eligible households in treatment villages that were enrolled in the program received a large, unconditional cash transfer of about USD 1,000 (nominal) in a series of three payments over eight months via the mobile money platform M-Pesa. These transfers were rolled out across study villages, with households beginning to receive transfers in 2014-15. A total of 10,500 households received transfers. The NGO identified 653 villages across three subcounties in Siaya County for potential expansion for their unconditional cash transfer program in 2014, covering over 65,000 households and approximately 280,000 individuals. These were all rural villages in the three subcounties that had not previously been a part of GD’s program. The study area was selected by the NGO due to its high poverty rates. In 2019, in preparation for a second endline survey, another household census was conducted and new households were incorporated into the sampling frame.

*Sampling Frame:* The sampling frame for Egger et al. (2019) was developed as follows: For our second endline, we augmented these 9,150 households with additional households identified in the 2019 census activity. We classify these new households into three groups: i) households that moved to the study area since our last census, ii) households that were newly formed from an existing household (most commonly children or siblings moving into their own household), iii) households that report having been in the study area at the time of our baseline census, but were not captured (this combines households that were actually present and missed with household misreporting). We include all households from group (ii) that split off of any of the 9,150 households in our original sample, as well as any split-offs from an additionally-drawn 18% of households not eligible for the cash transfer at baseline. In each village, we draw 24% of households belonging to groups (i) and (iii), but at least 1 household each. This provides a total sampling frame of 11,519 households.

**COVID-19 Survey Design:** Households targeted for inclusion in the second endline survey were included in two rounds of phone surveys, with the full sample targeted as part of each round.

*Sampling Frame:* Our total household sample for COVID-19 phone surveys includes 11,519 households that were planned for surveys as part of the second in-person endline surveys. *Survey Dates:* Round 1 of COVID phone surveys took place April 11–June 16, 2020. A second round was conducted June 18, 2020–September 2, 2020.

*Sample size, tracking and attrition:* 11,519 households.

*Median survey time:* 25 minutes.

*Sampling Weights:* Surveyed households are weighted by their 2019 census status in order to maintain population representativeness with our 2019 household census.

*IRB Approval:* UC Berkeley, Maseno University.

##### B.4.3 Kenya KEN3, Rural Households in Health Diaries project)

**Project Title:** Enhancing Universal Health Coverage in Kenya through Digital Innovations: A Financial and Health Diaries evaluation study of I-PUSH

**Target Population**: Rural households with pregnant women or children under 4 years old in Kakamega (Khwisero subcounty) and Kisumu (Kisumu East and Seme subcounties).

**Original Study Design:** Initial baseline data was collected in-person in October/November with weekly diaries (in-Person) anticipated from December 2019 for a duration of 12 months. The study would conclude with an endline survey after those 12 months. In Kakamega, the study design entailed a randomized control trial to evaluate the impact of a mobile phone-based health insurance scheme. In Kisumu, the study was designed to evaluate free access to public care through a prospective longitudinal analysis.

*Sampling Frame:* The study population, drawn from low-income rural villages, consists of households with either a pregnant woman or a mother with children below four years old. First, 32 villages were randomly selected from the catchment areas of six health facilities, 24 in Kakamega and 8 in Kisumu. Next, in each village in Kakamega, ten households were randomly sampled from lists with households fulfilling the study eligibility criteria, resulting in 240 sampled households. In Kisumu, between 11 and 15 households per village were randomly selected resulting in 107 sampled households.

**COVID-19 Survey Design:** Phone surveys. After the onset of the first COVID-19 case in Kenya (mid-March 2020), the weekly diaries data collection transitioned from in-person interviewing to phone interviews.

*Sampling Frame:* Since weekly data collection was ongoing, the sampling frame for the phone surveys was the same as for the original study. In addition to the original survey modules, a special COVID-19 module was added in May 2020, including questions on symptoms of COVID-19, changes in health care utilization, and knowledge, attitudes and behaviours in response to COVID-19 preventive measures.

*Survey Dates:* October 18, 2019 to July 15, 2021

*Sample size, tracking and attrition:* Kakamega: 240 households, of which 44 dropped out between baseline and midline, while 57 were added to the sample, resulting in 253 households at the midline point. Kisumu: 107 households, of which 18 dropped out between baseline and midline, while 4 were added, resulting in a midline sample of 93 households.

*Median survey time:* N/A.

*Sampling Weights:* None.

*IRB Approval:* AMREF Ethics and Scientific Review Committee Kenya P679/2019 (August 8, 2019) with an extension granted on April 21, 2020.

#### B.5 Nepal

##### COVID-19 Experience

*Case History:*

- First COVID-19 case confirmed on January 23, 2020
- Second case confirmed on March 23, 2020
- Total cases: 662,570 as of July 15, 2021 (Our World in Data)
- Total deaths: 9,463 as of July 15, 2021 (Our World in Data)

*Mobility Restrictions:*

- January 28, 2020: Land border with China closed
- March 22, 2020: All international flights stopped
- March 23, 2020: Land border with India closed
- March 24, 2020: National lockdown takes effect. Movement outside the home banned except to purchase necessities or receive medical care. Motorized vehicles without prior permission banned from use. All transport services banned. Extended to July 21, 2020.
- August 11, 2020: Only passengers that have permission from the District Covid Crisis Management Centre and Local Administration will be allowed to travel from one district to another.
- September 21, 2020: Domestic passenger flights and long-distance public transport has resumed. International flights have been allowed from October 1, 2020.

*Social Distancing:*

- March 18, 2020: Gatherings of more than 25 people banned, including places of worship.
- August 11, 2020: Permission for daily prayers festivals observed among family members at home.

*School Closures:*

- March 19, 2020 - September 17, 2021: All classes and examinations suspended.

*Social Protection Responses:*

- The government provided food aid packages to an unspecified number of vulnerable households, with one source estimating that 70 - 95% of households in this category had received assistance (Franciscon & Arruda 2020).
- Plans for utility fee waivers and a public works program for individuals in the informal sector were announced, although there is limited data on how many people have benefitted from these schemes (Gentilini et al. 2020).

##### Economic Context

*Seasonality and Food Security* In 2019, Nepal produced record-level cereal crops, the latest in a series of four consecutive bumper harvests. The Nepal post-COVID survey was conducted several months before the start of the 2020 lean period. However, the FAO remained concerned about food insecurity for approximately 15% of the Nepalese population. (Global Information and Early Warning System on Food and Agriculture Country Brief: Nepal, *Food and Agricultural Organisation of the United Nations*, 13-May-2020).

*Social Protection* Nepal has over 80 social protection schemes in operation (Ghimire 2019), with more than half of their spending going towards people who are elderly or disabled, and the rest distributed between a variety of employment and scholarship schemes (Sijapati 2017). While coverage is fairly high, at 28% of the population, the average program provides very low levels of benefits, at only $2 - $5 per month (Sijapati 2017).

##### B.5.1 Nepal NPL, Agricultural Households in Western Terai

**Project Title:** Western Terai Panel Survey (WTPS)

**Target Population**: Rural households in the districts of Kailali and Kanchanpur.

**Original Study Design:** Initial baseline data was collected in-person in July of 2019, and 5 rounds of phone survey data were collected between August 12, 2019 and January 4, 2020. *Sampling Frame:* The phone survey sample includes 2,636 rural households in the districts of Kailali and Kanchanpur, which represent the set of households that responded to phone surveys from an original sample of 2,935 households. This sample was constructed by randomly sampling 33 wards from 15 of the 20 sub-districts in Kanchanpur and selecting a random 97 villages from within those wards. At the time of baseline data collection in July of 2019, 7 of these 97 villages were dropped from the sample due to flooding. Households belong to the bottom half of the wealth distribution in these villages, as estimated by a participatory wealth ranking exercise with members of the village.

**COVID-19 Survey Design:** Phone surveys

*Sampling Frame:* Two phone surveys were fielded in April, 2020. The first included detailed questions on social distancing and more sparse data on other socioeconomic variables. This survey attempted to reach the universe of 1,820 households that had responded to at least one prior phone call in 48 villages, and successfully reached 1,419 households. The second survey included only sparse information on social distancing and more detailed questions on socioeconomic variables. This survey attempted to reach a random sample of 500 households across the remaining 42 villages, and successfully reached 408.

*Survey Dates:* April 1 to April 29, 2020

*Sample size, tracking and attrition:* 1,981 households

*Median Survey Time:* 27 minutes (COVID-19 survey) and 18 minutes (Socioeconomic survey)

*Sampling Weights:* None.

*IRB Approval:* Yale University IRB Protocol 2000025621

#### B.6 Nigeria

##### COVID-19 Experience

*Case History:*

- First confirmed case: February 27, 2020
- Total cases: 169,074 as of July 15, 2021 (Our World in Data)
- Total deaths: 2,126 as of July 15, 2021 (Our World in Data)

*Mobility Restrictions:*

- March 18, 2020: Travel ban on hot-spot countries.
- March 23-September 5, 2020: International flights banned.
- March 29, 2020 - May 18, 2020: Partial lockdowns in hot-spot states.
- April 20 - June 8, 2020: Domestic flights banned.
- April 27, 2020 - May 7, 2020: Nationwide ban on interstate movement.
- May 2, 2020: Nationwide overnight curfew. Revised to 12am-4am in September 3, 2020.
- June 2, 2020: Reopening of worship centers.
- July 1, 2020: Lifting of nationwide travel ban.

*School Closures:*

- March 26, 2020: Closure of all schools and higher education institutions.
- July 1, 2020: School resumption for graduating students.
- September 5, 2020: School resumption nationwide with COVID-19 guidelines in place.

*Social Distancing:*

- March 18, 2020: Gatherings of more than 50 people banned in hot-spot states.
- April 27, 2020: Mandatory use of face masks in public.

*Social Protection Responses:*

- March 23, 2020: Central Bank of Nigeria announced stimulus package for households, SMEs and health sector.
- March 27, 2020: Government reached out private donors to raise $330 billion.
- April 1, 2020: Federal Ministry of Humanitarian Affairs, Disaster Management and Social Development (FMHADMSD) announced free food rations to needy.
- April 1, 2020: Government announcement on cash transfer ($52) to poor registered in National Social Register.
- Arpil 6, 2020: Government reached out multilateral donors to raise $6.9 billion.
- May 7, 2020: Government raised $4.34 billion from domestic stock market to finance the budget.

##### Economic Context

*Seasonality* The NGA pre-COVID survey was conducted several months before the start of the 2020 lean period. However, the Boko Haram insurgency has led to heightened levels of displacement and food insecurity in the region. Nigeria alone accounts for 42% of the region’s total number of acutely food-insecure people. In terms of absolute numbers, Nigeria ranked among the world’s 10 worst food crises in 2019, with 5 million food insecure people. (Food and Nutrition Crisis 2020, Analyses Responses, Maps Facts, *SWAC/OECD*, 2020).

*Social Protection* The Federal Government led social protection includes the following programmes:

- The National Social Safety Net Project (NASSP)
- In-Care of the Poor (COPE), which was funded initially through the MDGs-DRG fund, targeted at extremely poor households (those headed by a female, the elderly, physically challenged,fistula or HIV/AIDS patients) with children of school-going age;
- The health fee waiver for pregnant women and under-fives (funded by the MDGs-DRG and provided on a universal basis)
- The Community-Based Health Insurance Scheme (CBHIS) (re-launched in 2011)
- SURE - P MNCH, CSWYE programmes funded from the savings from the oil subsidy reform in 2013
- At the State level, social protection programmes cover a range of broad interventions, which are implemented by government Ministries, Departments and Agencies (MDAs) and/or funded by international donors.

However, only about 2% of Nigerians were enrolled in the country’s major social security program, the National Social Safety Net Project (NASSP), and most other programs had significantly lower coverage. Throughout the COVID-19 crisis, coverage of social safety services remained low. Between March 2020 and March 2021, only 4% of households received federal, state, or municipal government assistance in the form of cash (Jonathan Lain Tara Vishwanath, 2021).

##### B.6.1 Nigeria NGA, Young Adults in Edo and Delta States, Nigeria

**Project Title:** Irregular Migration and Misinformation in Nigeria

**Target Population**: Young adults in Edo and Delta states, Nigeria

**Original Study Design:** Initial baseline data was collected in-person from March 9 to 23, 2020, followed by a Covid-19 phone survey carried out from May 20 to July 23, 2020. Follow-up data for this sample was collected by way of a phone survey from April 22 to May 21, 2021. Respondents that could not be reached by phone received an in-person follow-up visit between August 9 and September 1, 2021.

*Sampling Frame:* We georeferenced all residential structures in the states of Edo and Delta states from satellite imagery, drew enumeration areas, and randomly selected enumeration areas and structures for inclusion in our sample. Enumerators then located structures using GPS-enabled devices, and randomly selected households within buildings. After obtaining the head of household’s consent, enumerators completed a brief household survey module and roster using a tablet, and the device randomly selected an eligible individual from each household roster for an in-depth interview. Eligible individuals included all resident men and women aged 18 to 39.

**COVID-19 Survey Design:** Phone survey.

*Sampling Frame:* For the Covid-19 phone survey, we attempted to reach all individuals with whom a baseline interview had been conducted in March 2020.

*Survey Dates:* The Covid-19 phone survey took place from May 20 to July 23, 2020. Follow-up mental health data was collected by phone from April 22 to May 21, 2021, and in person from August 9 to September 1, 2021.

*Sample size, tracking and attrition:* Follow-up data is available for 628 individuals, out of a relevant baseline sample of 880 individuals.

*Median Survey Time:* The baseline survey included a 15-minute household module and roster and a 60-minute in-depth individual interview. The Covid-19 and follow-up surveys typically required 30 minutes.

*Sampling Weights:* None.

*IRB Approval:* IPA IRB Protocol 15057.

#### B.7 Rwanda

##### COVID-19 Experience

*Case History:*

- First confirmed case: March 14, 2020
- Total cases: 51,625 as of July 15, 2021 (Our World in Data)
- Total deaths: 616 as of July 15, 2021 (Our World in Data)

*Mobility Restrictions:*

- March 18, 2020: All commercial passenger flights halted.
- March 21, 2020: Travel between different cities and districts prohibited except for medical reasons or essential services. Borders closed except for goods and cargo and returning Rwandan citizens and legal residents.
- March 21, 2020 - May 4, 2020: Nationwide lockdown. Movement for essential services allowed.
- April 25, 2020: People in need of essential services must request clearance online and wait for approval before attempting movement.
- May 4, 2020: Curfew instituted between 8 pm to 5 am. Curfew changed to 9 pm to 5 am on May 18 and to 10 pm to 4 am on Oct 13, 2020 until Nov, 2021.
- June 2, 2020: Travel between provinces is allowed.
- July 8, 2020: Public and private businesses reopened with essential staff.

*School Closures:*

- March 16, 2020: Schools closed for four weeks. On April 30, 2020, fully opened in February 2021.

*Social Distancing:*

- March 8, 2020: Concerts and other public gatherings that bring many people together postponed.
- March 15, 2020: Places of worship closed.
- April 19, 2020: Face masks required in public and in multi-family compounds.
- April 30, 2020: Meetings in public and mass gatherings prohibited.
- June 16, 2020: Domestic tourism and international tourism for visitors traveling with charter flights resumed.
- June 30, 2020: Non-contact outdoor sports permitted.
- July 8, 2020: Religious weddings, burials and funeral gatherings are allowed with no more than 30 people.
- August 14, 2020: Public and private sector offices allowed to work in person with 50% capacity.

*Social Protection Responses:*

- March 28, 2020: Food distributions reaching 20,000 beneficiaries initiated in three districts of Kigali, starting with urban poor who cannot work and have no garden. Government fixed prices for 17 basic food items. (Global Information and Early Warning System on Food and Agriculture Country Brief: Rwanda, *Food and Agricultural Organisation of the United Nations*, 19-June-2020)
- May 4, 2020: People able to resume work will no longer receive food.
- Government added 56,000 new families to its existing Vision 2020 Umurenge Program (VUP) social protection scheme (Gentilini et al. 2020)

##### Economic Context

*Seasonality* The post-COVID survey was conducted during the lean season. The first 2020 harvest produced above-average yields. Flooding and landslides earlier in 2020 did not impact the first harvest and minimally impacted the anticipated second harvest. After exceptionally high bean and maize prices in December 2019, prices declined through February with the first harvest, increased during early pandemic closures, and then began to decrease again in May 2020 (Global Information and Early Warning System on Food and Agriculture Country Brief: Rwanda, *Food and Agricultural Organisation of the United Nations*, 19-June-2020).

*Social Protection* Rwanda runs a variety of social protection programs, including public works and cash transfer programs, under the umbrella of its Vision 2020 Umurenge Program (VUP). Over a million people have benefitted from this program (World Bank 2019), approximately 7.5% of the population (Beegle, Coudousel & Monsalve 2018).

##### B.7.1 Rwanda RWA, Participants in 100WEEKS cash tranfer program

**Project Title:** 100WEEKS impact measurement.

**Target Population**: Women between 20-40 years of age living in poverty in Musanze, Rwanda.

**Original Study Design:** Baseline interview, 5 phone interviews, endline interview after 100 weeks.

*Sampling Frame:* Participants have been surveyed as part of the monitoring activities of a cash transfer program. The target group for the implementation of the cash transfer program were women between 20-45 years of age, who are in the lowest income quintile and are expected to have the ability to move out of poverty.

**COVID-19 Survey Design:** Phone surveys.

*Sampling Frame:* The sampling frame is the same as the original study design. Phone surveys were already a standard part of the monitoring efforts of 100WEEKS. The face-to-face baseline interviews that are conducted at baseline were postponed during the 2-month lockdown in Rwanda, which meant the newly selected women had to wait to start the program.

*Survey Dates:* 25 June 2016 - 01 April 2021.

*Sample size, tracking and attrition:* 843 women were drop out.

*Median Survey Time:* For the face-to-face interviews the average survey time is 45 minutes, depending on the experience of the enumerator. The phone interviews take 10-25 minutes, depending on the survey round.

*Sampling Weights:* None.

*IRB Approval:* Vrije University Amsterdam - 20210413.1.wjs400 (June 7, 2021)

#### B.8 Sierra Leone

##### COVID-19 Experience

*Case History:*

- First confirmed case: March 31, 2020
- Total cases: 6,122 as of July 15, 2021 (Our World in Data)
- Total deaths: 113 as of July 15, 2021 (Our World in Data)

*Mobility Restrictions:*

- March 16, 2020: Imposed quarantine on all international travelers. Non-essential government and businesses directed to begin working from home.
- March 22, 2020: National and international flights halted.
- April 5, 2020 - April 11, 2020 : Lock down 1
- April 11, 2020: Start of dusk to dawn curfew (9pm-6am). Changed to 11pm-6am on June 24, 2020, and 11pm-5am on July 3, 2020. Inter-District travel banned.
- May 3-5, 2020: Lock down 2
- June 23, 2020: Inter-District travel restrictions lifted
- July 13, 2020: All worship gatherings were allowed.

*Social Distancing:*

- April 7, 2020: Limits on large gatherings, church attendance, closure of courts; Safari-com suspends mobile money transaction fees to encourage cashless transactions
- June 23, 2020: Mask wearing in public spaces mandatory

*School Closures:*

- March 31, 2020 - October 5, 2020: Closure of all schools and higher education institutions.

*Social Protection Responses:*

- April 19-21, 2020: 25 USD Cash transfers and 25 Kgs of rice to Persons with Disabilities during Lockdown 1 (April 2020)
- May 3-5, 2020: 25 kgs of rice to persons with disabilities during Lockdown 2 (May 2020)
- Emergency cash transfer for 29,000 petty traders starting in June 2020. They have also increased the value of the cash transfers provided to existing beneficiaries of the safety net program (Gentilini et al. 2020). Estimates of the coverage of these cash transfers varies. Gentilini et al. (2020) suggest that up to 14% of the population should benefit from these programs
- Lower-interest loans and tax deferments for SMEs, and cash transfers to vulnerable female heads of households.

##### Economic Context

*Seasonality* The post-COVID surveys were conducted during the lean season. The FAO had estimated that one million people would require food aid between March and May 2020, and in its May 2020 report assessed that 1.3 million could need food aid during the June-August 2020 period absent mitigation efforts. (Global Information and Early Warning System on Food and Agriculture Country Brief: Sierra Leone, *Food and Agricultural Organisation of the United Nations*, 05-May-2020).

*Social Protection* Sierra Leone launched its Social Safety Net program to support vulnerable households in 2013 (FAO 2019). Coverage of this program has been quite limited, reaching only 2.3% of the population (Beegle, Coudousel & Monsalve 2018).

##### B.8.1 Sierra Leone (SLE), Participants in the Sierra Leone Rural Electrification project

**Project Title:** Sierra Leone Rural Electrification (SLRE)

**Target Population**: Households in 195 rural towns across all 12 districts of Sierra Leone. Of these, 97 villages were selected to benefit from an electrification program.

##### Original Study Design

*Intervention:* The Government of Sierra Leone (GoSL) in collaboration with United Nations Office for Project Services (UNOPS) and international donors is implementing the Rural Renewable Energy Project (RREP). In its first wave, during 2017, the project provided stand-alone solar photovoltaic powered mini-grids to 54 communities across the country. Construction of mini-grids in a further 43 towns is ongoing. In selected communities, engineers construct 6kW–36kW power mini-grids that provide reliable power year-round. Electricity is free for schools and clinics. Residential and commercial users can acquire connections from commercial operators.

*Sampling Frame:* Household data was collected in 195 towns across all 12 districts of Sierra Leone. The GoSL selected 97 towns with (planned) mini-grids. We use Propensity Score Matching to select 98 control communities. Within communities, respondents were randomly selected from a census roster stratified by occupation status of farmers, business owners and other occupations [47 percent, 47 percent and 7 percent]. In each village, the intended sample was 43 households (20 farmers, 20 business, 3 other). Data was collected during June–July (108 communities) and November–December 2019 (87 communities). If a household on our sampling list was not available on the village visit day, we had a randomly sampled list of replacement households to survey. The replacement household would be the same occupation as the sampled household would have been so the sample ratio of 20-20-3 still held in each community. Total baseline sample constitutes 3,230 respondents.

**COVID-19 Survey Design:** An additional round of survey data was collected during March - April 2021, in the same communities and under the same households. Total followup survey sample constitutes 2,808 respondents in 108 villages.

*Sampling Frame:* See above.

*Survey Dates:* First round data was collected during June–July (108 communities) and November–December 2019 (87 communities). An additional round of survey data was collected during March - April 2021, in the same communities and under the same households. *Sample size, tracking and attrition:* Total baseline sample constitutes 3,230 respondents, total followup survey sample constitutes 2,808 respondents. Attrition rate is 13%.

*Median survey time:* 33 minutes.

*Sampling Weights:* None.

*IRB Approval:* Sierra Leone Ethics and Scientific Review Committee (SLERC 2904202) and Wageningen University (24062020).

### C. Research approvals

**BGD. Rohingya refugees from Myanmar: Refugee camp households in Bangladesh**: BRAC University (ref no. 2019-028-ER).

**COL. Primary caregivers in Tumaco, Colombia**: the Ethics Committee of the Universidad de los Andes, Colombia (protocol 786, 2017).

**DRC. Impact evaluation of Community Based Trauma Healing)**: the NORC Institutional Review Board (IRB00000967, IRB Protocol Number 18.08.13).

**KEN1. Participants in child health and human capital intervention in Kenya**: UC Berkeley, Maseno University.

**KEN2. Rural Households in NGO Cash Transfer Study**: UC Berkeley, Maseno University.

**KEN3. Rural Households in Health Diaries project**: AMREF Ethics and Scientific Review Committee Kenya P679/2019.

**NPL. Agricultural Households in Western Terai**: Yale University IRB Protocol 2000025621.

**NGA. Young Adults in Edo and Delta States, Nigeria**: IPA IRB Protocol 15057.

**RWA. Participants in 100WEEKS cash tranfer program**: Vrije University Amsterdam (20210413.1.wjs400).

**SLE. Participants in the Sierra Leone Rural Electrification project**: Sierra Leone Ethics and Scientific Review Committee (SLERC 2904202) and Wageningen University (24062020).

## Footnotes

1 In some cases where very few interviews were conducted in the final month of a survey wave and the interview dates do not span the entire final month, we merge the final two months of the survey wave into one time period that spans between one and two months.

2 The weights applied to these samples in the random-effects aggregate are 20, 13, 25, 15, 22, and 6, respectively, for our preferred depression index. Weights are proportional to 1*/*((*SEs*)^2^ + *tau*^2^) where *SEs* is the standard error of the estimate in sample *s*, and *tau*^2^ is our estimate of the variance in “true effects” across countries. RWA is weighted low due to its particularly large confidence intervals.

3 We omit one eligible sample from this analysis (KEN3) because – unlike the other samples – we are uncertain about the most appropriate model for estimating long-run impacts for this sample, while its long-run results are highly sensitive to model assumptions. The weights for each sample in average are 13, 11, 13, 8, 12, 12, 13, 13, and 7, respectively. Weights are calculated as for the short-run aggregate.

